# Life-years lost associated with mental illness: a cohort study of beneficiaries of a South African medical insurance scheme

**DOI:** 10.1101/2023.01.19.23284778

**Authors:** Yann Ruffieux, Anja Wettstein, Gary Maartens, Naomi Folb, Cristina Mesa Vieira, Christiane Didden, Mpho Tlali, Chanwyn Williams, Morna Cornell, Michael Schomaker, Leigh F Johnson, John A Joska, Matthias Egger, Andreas D Haas

**Author notes:** Corresponding author: Andreas D. Haas, Mittelstrasse 43, 3012 Bern, Switzerland. Joint first authors (Yann Ruffieux, Anja Wettstein).

## Abstract

**Importance:** People with mental illness have a reduced life expectancy, but the extent of the mortality gap and the contribution of natural and unnatural causes to excess mortality among people with mental illness in South Africa are unknown.

**Objective:** To quantify excess mortality due to natural and unnatural causes associated with mental illness.

**Design, setting and participants:** Cohort study using reimbursement claims and vital registration of beneficiaries of a South African medical insurance scheme, aged 15-84 years and covered by medical insurance at any point between January 1, 2011, and June 30, 2020.

**Exposures:** ICD-10 diagnoses of mental disorders including organic, substance use, psychotic, mood, anxiety, eating, personality, and developmental disorders.

**Outcomes:** Mortality from natural, unnatural, unknown and all causes, as measured by the life-years lost (LYL) metric.

**Results:** We followed 1 070 183 beneficiaries (51.7% female, median age 36.1 years for a median duration of 3.0 years, of whom 282 926 (26.4%) received mental health diagnoses and 27 640 (2.6%) died. Life expectancy of people with mental health diagnoses was 3.83 years (95% CI 3.58-4.10) shorter for men and 2.19 years (1.97-2.41) shorter for women. Excess mortality varied by sex and diagnosis, ranging from 11.50 LYL (95% CI 9.79-13.07) among men with alcohol use disorder to 0.87 LYL (0.40-1.43) among women with generalised anxiety disorder. Most LYL were attributable to natural causes (3.42 among men and 1.94 among women). A considerable number of LYL were attributable to unnatural causes among men with bipolar (1.52) or substance use (2.45) disorder.

**Conclusions and Relevance:** The burden of premature mortality among persons with mental disorders in South Africa is high. Our findings support implementing interventions for prevention, early detection, and treatment of physical comorbidities among people with mental disorders. Suicide prevention and substance use treatment programmes are needed to reduce excess mortality from unnatural causes, especially among men.

**Key points:** *Question:* How much shorter is the life expectancy of people with mental illness compared to the general population and how many life years are lost due to natural and unnatural causes of death?

*Findings:* The life expectancy of people with mental health diagnoses was 3.83 years shorter for men and 2.19 years shorter for women. Most excess life years lost were attributable to natural causes (3.42 among men and 1.94 among women). However, bipolar and substance use disorders were associated with considerable premature mortality from unnatural causes.

*Meaning:* Our findings support the implementation of interventions for improving the physical health of people with mental illness and targeted suicide prevention and substance use treatment programmes.

## Introduction

Mental illness is a leading and increasing cause of disease burden globally.^1^ In South Africa, the total number of disability-adjusted life years attributable to mental illness nearly doubled from 550 000 to 929 000 over the past 30 years.^1^ Mental disorders now rank among the top ten leading causes of disease burden in South Africa,^1^ with a lifetime prevalence of mental disorders in South Africa estimated at 30%.^2^

Although most mental disorders do not lead directly to death, they increase the risk of suicide, accidental death, and premature mortality from physical illness.^3–6^ Higher rates of premature mortality from physical illness can be attributed to a higher incidence of physical comorbidities among people with mental illness and worse access to or engagement in health care.^4,7^ Physical illnesses can also increase the risk of mental illness due to various biological and psycho-social mechanisms.^8^

Meta-analyses of studies—mainly from high-income countries—demonstrated that people with mental illnesses, particularly those with severe conditions, have more than double the mortality rates of the general population and die on average 10 years prematurely.^5,6^ Due to differences in life expectancy and disease burden, estimates from high-income countries cannot be generalized to low- and middle-income settings. However, evidence from low- and middle-income settings^5,6^ is scarce.^9–11^

We aimed to investigate excess mortality among people with mental illness in South Africa. We analyzed longitudinal reimbursement claims and vital registration data of beneficiaries of a large South African medical insurance scheme. We calculated excess life years lost (LYL) and adjusted hazard ratios (HRs) for excess mortality among people diagnosed with a mental illness.

## Methods

### Study design and participants

We analyzed longitudinal reimbursement claims and vital registration data of a cohort of beneficiaries of a large South African medical insurance scheme. Reimbursement claims contained International Classification of Diseases, tenth revision (ICD-10) diagnoses made in outpatient and hospital settings. Eligible beneficiaries for our analysis were aged 15–84 years and covered by medical insurance at any point between January 1, 2011, and June 30, 2020. We excluded people of unknown age and sex and those whose vital status could not be ascertained based on the linkage to the National Population Register (NPR) (appendix, figure S1). The Human Research Ethics Committee of the University of Cape Town and the Cantonal Ethics Committee of the Canton of Bern granted permission to analyze these data.

### Procedures

Our primary exposures were ICD-10 diagnoses of mental disorders made in outpatient or hospital settings. We grouped diagnoses into organic mental disorders (ICD-10 codes F00-F09), substance use disorders (F10–F17, F19), psychotic disorders (F20–F29), mood disorders (F30–F39), anxiety disorders (F40–F48), eating disorders (F50), personality disorders (F60–F69), developmental disorders (F80–F89), or any mental disorder (F00–F99). We further subdivided substance use disorders into alcohol use disorder (F10) and drug use disorders (F11–F17, F19), mood disorders into bipolar disorders (F31) and depressive disorders (F32–F33, F34.1), and anxiety disorders into generalized anxiety disorder (F41.1) and post-traumatic stress disorder (F43.1).

In secondary analyses, we only analyzed ICD-10 diagnoses made in a hospital setting and psychiatric medication prescriptions as proxies for mental disorders. We grouped psychiatric medication following the World Health Organization’s Anatomical Therapeutic Chemical classification system into substance use medication (N07B), antipsychotic (N05A), antidepressant (N06A), anxiolytic (N05B), or any psychiatric medication (N05A, NO5B, N06A, and N07B).

We ascertained the vital status of beneficiaries based on mortality records from NPR and the medical insurance database. If the death dates recorded by the medical insurance did not match NPR data, we used death dates from NPR. Finally, we divided mortality by natural, unnatural, or unknown causes of death. Unnatural causes include all deaths by external causes (ICD10 V01—Y98) and natural causes include all causes of death from chapters 1 to 18 of the ICD-10.^12^ Unknown causes include death under investigation at the time of linkage, death due to unidentified causes, and death recorded only by the medical insurance but not by the NPR.

### Statistical analysis

For each individual, we defined baseline as either the date of enrolment in the medical insurance scheme, their 15^th^ birthday, or January 1, 2011, whichever occurred later. We followed people from baseline to the end of insurance coverage, death, their 85^th^ birthday, or June 30, 2020, whichever occurred first. We assessed the distribution of sex, baseline age, and baseline calendar year for participants with and without mental health diagnoses.

We estimated LYL of people with mental health diagnoses and psychiatric medication. LYL measures the average difference in life expectancy among people with mental health diagnoses between the dates of diagnoses and their 85^th^ birthday, compared to people of the same age and sex without mental health diagnoses.^13–16^ Individuals were considered unexposed until receiving a first mental health diagnosis and exposed from the date of their first diagnosis of the mental illness of interest. We disaggregated LYL by cause of death (natural, unnatural, or unknown).^17^ We produced 95% CI for the LYL estimates using bootstrap simulation. In sensitivity analyses, we restricted analysis of LYL to persons with a single ICD-10 diagnosis of a mental disorder that could result from a coding error. In another sensitivity analysis, we did not censor follow-up at the end of insurance coverage if the person died within 1 year of the end of insurance coverage.

Using multivariable Cox proportional hazard models, we estimated HRs with 95% CIs for associations between each exposure and mortality from all causes, natural causes, and unnatural causes. All models were adjusted for the exposure of interest (time-updated variable with the individual switching from unexposed to exposed at the time of the first diagnosis of the mental illness), baseline age (categories 15–24, 25–39, 40–54, 55–74, 75–84), and calendar period (time-updated by categories January 1, 2011– December 31, 2013; January 1, 2014–December 31, 2016; January 1, 2017–March 14, 2020; March 15– June 30, 2020). We fitted separate models adjusting and not adjusting for psychiatric comorbidity. We tested for interactions between sex and each exposure. We estimated HRs for each sex and with both sexes combined while adjusting for sex. We performed additional analyses where we split analysis time at one- and two-year marks after the start of time-at-risk and produced an adjusted HR for each interval.

We performed statistical analyses using R 4.1.2 (R Foundation for Statistical Computing, Vienna, Austria) and the *lillies* package.^16^

## Results

We followed 1 070 183 people (517 305 men and 552 878 women) for a median duration of 3.0 years [interquartile range (IQR) 1.2–6.1) (appendix, Figure S1). The characteristics of people with and without mental health diagnoses are shown in Table 1.

**Table 1:**
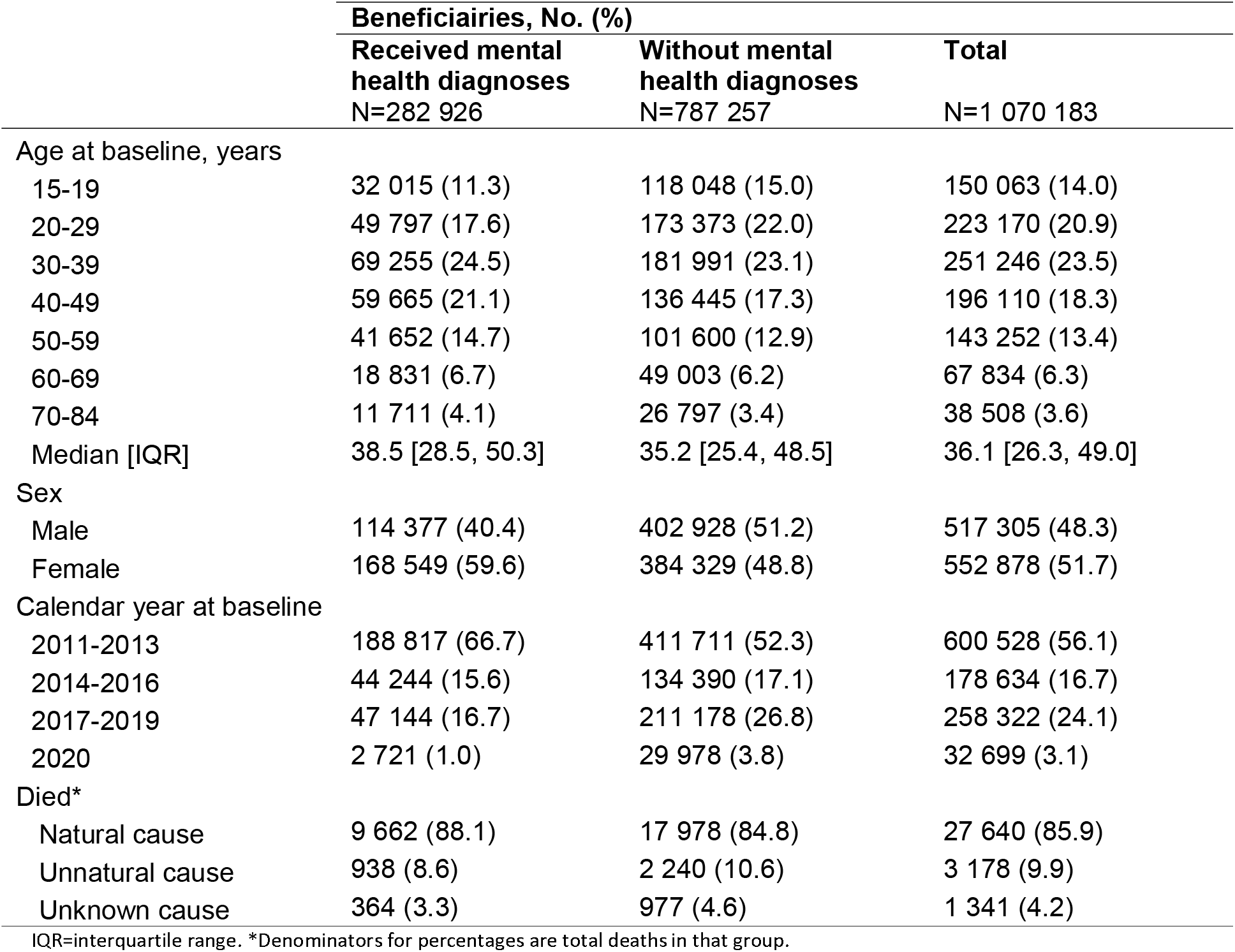
Characteristics of people who did and did not receive mental health diagnoses before their end of follow-up

|  | Beneficiaries, No. (%) |  |  |
| --- | --- | --- | --- |
|  | Received mental health diagnoses<br>N=282 926 | Without mental health diagnoses<br>N=787 257 | Total<br>N=1 070 183 |
| Age at baseline, years |  |  |  |
| 15-19 | 32 015 (11.3) | 118 048 (15.0) | 150 063 (14.0) |
| 20-29 | 49 797 (17.6) | 173 373 (22.0) | 223 170 (20.9) |
| 30-39 | 69 255 (24.5) | 181 991 (23.1) | 251 246 (23.5) |
| 40-49 | 59 665 (21.1) | 136 445 (17.3) | 196 110 (18.3) |
| 50-59 | 41 652 (14.7) | 101 600 (12.9) | 143 252 (13.4) |
| 60-69 | 18 831 (6.7) | 49 003 (6.2) | 67 834 (6.3) |
| 70-84 | 11 711 (4.1) | 26 797 (3.4) | 38 508 (3.6) |
| Median [IQR] | 38.5 [28.5, 50.3] | 35.2 [25.4, 48.5] | 36.1 [26.3, 49.0] |
| Sex |  |  |  |
| Male | 114 377 (40.4) | 402 928 (51.2) | 517 305 (48.3) |
| Female | 168 549 (59.6) | 384 329 (48.8) | 552 878 (51.7) |
| Calendar year at baseline |  |  |  |
| 2011-2013 | 188 817 (66.7) | 411 711 (52.3) | 600 528 (56.1) |
| 2014-2016 | 44 244 (15.6) | 134 390 (17.1) | 178 634 (16.7) |
| 2017-2019 | 47 144 (16.7) | 211 178 (26.8) | 258 322 (24.1) |
| 2020 | 2 721 (1.0) | 29 978 (3.8) | 32 699 (3.1) |
| Died* |  |  |  |
| Natural cause | 9 662 (88.1) | 17 978 (84.8) | 27 640 (85.9) |
| Unnatural cause | 938 (8.6) | 2 240 (10.6) | 3 178 (9.9) |
| Unknown cause | 364 (3.3) | 977 (4.6) | 1 341 (4.2) |

IQR=interquartile range. \*Denominators for percentages are total deaths in that group.

During follow-up, 282 926 beneficiaries (26.4%) received a mental health diagnosis (Table 2). The proportion of beneficiaries who received mental health diagnoses was higher among women (30.5%) than men (22.1%). The most common mental health diagnoses were anxiety disorders (16.6%) and mood disorders (14.6%). The proportion of beneficiaries diagnosed with organic, substance use, psychotic, eating, developmental, and personality disorders was below 1% (Table 2). Psychiatric comorbidity was common. For example, among beneficiaries diagnosed with a psychotic disorder, 75.2% had also been diagnosed with a mood disorder (appendix, Figure S2). A total of 45 579 beneficiaries (4.3%) received a mental health diagnosis in a hospital (appendix, Table S1). Over one-third of the study population was prescribed a psychiatric medication (34.8%) (appendix, Table S2).

**Table 2:** Numbers and proportions of people who received mental health diagnoses during follow-up by sex

|  | **Beneficiaries, No. (%)** |  |  |
| --- | --- | --- | --- |
|  | **Men**<br>N=517 305 | **Women**<br>N=552 878 | **Total**<br>N=1 070 183 |
| Any Mental Health Diagnosis (F00-F99) | 114 377 (22.1) | 168 549 (30.5) | 282 926 (26.4) |
| Organic Mental Disorders (F00-F09) | 3 863 (0.7) | 4 655 (0.8) | 8 518 (0.8) |
| Substance Use Disorders (F10-F17, F19) | 5 275 (1.0) | 2 042 (0.4) | 7 317 (0.7) |
| Alcohol Use Disorders (F10) | 2 757 (0.5) | 938 (0.2) | 3 695 (0.3) |
| Drug Use Disorders (F11-F17, F19) | 2 947 (0.6) | 1 226 (0.2) | 4 173 (0.4) |
| Psychotic Disorders (F20-F29) | 2 228 (0.4) | 2 324 (0.4) | 4 552 (0.4) |
| Mood Disorders (F30-F39) | 57 038 (11.0) | 98 835 (17.9) | 155 873 (14.6) |
| Bipolar Disorders (F31) | 6 786 (1.3) | 12 078 (2.2) | 18 864 (1.8) |
| Depression (F32-F33, F34.1) | 54 755 (10.6) | 95 785 (17.3) | 150 540 (14.1) |
| Anxiety Disorders (F40-F48) | 67 599 (13.1) | 110 405 (20.0) | 178 004 (16.6) |
| Generalized Anxiety Disorders (F41.1) | 11 521 (2.2) | 21 062 (3.8) | 32 583 (3.0) |
| Post-traumatic Stress Disorders (F43.1) | 5 362 (1.0) | 8 593 (1.6) | 13 955 (1.3) |
| Developmental Disorders (F80-F89) | 2 353 (0.5) | 1 646 (0.3) | 3 999 (0.4) |
| Eating Disorders (F51) | 459 (0.1) | 897 (0.2) | 1 356 (0.1) |
| Personality Disorders (F60-F69) | 653 (0.1) | 752 (0.1) | 1 405 (0.1) |

International Classification of Diseases, tenth revision code range of each diagnostic category is shown in parenthesis.

Figure 1 shows LYL among men and women who received mental health diagnoses in any health care setting (left panel) or hospital (right panel) compared to beneficiaries of the same sex and age without mental health diagnoses. Men had higher LYL than women for all types of mental illnesses except developmental disorders. For beneficiaries diagnosed in any health care setting, life expectancy after diagnosis of any mental disorder was 3.83 years (95% CI 3.58–4.10) shorter for men and 2.19 years (1.97–2.41) shorter for women when compared to beneficiaries of the same age and sex without mental health diagnoses. The life expectancy of men diagnosed with an organic mental disorder, alcohol use disorder, drug use disorder, or psychotic disorder was reduced by over 10 years. Women with organic or alcohol use disorder had a similar mortality gap of 10 years. Psychotic and drug use disorders among women and developmental and eating disorders for both sexes were associated with an LYL of 7 to 10 years. Women with common mental disorders showed low excess mortality, for example: 0.87 (95% CI 0.40–1.43) LYL for generalized anxiety disorder and 2.47 (2.17–2.74) LYL for depressive disorders.

**Figure 1:**
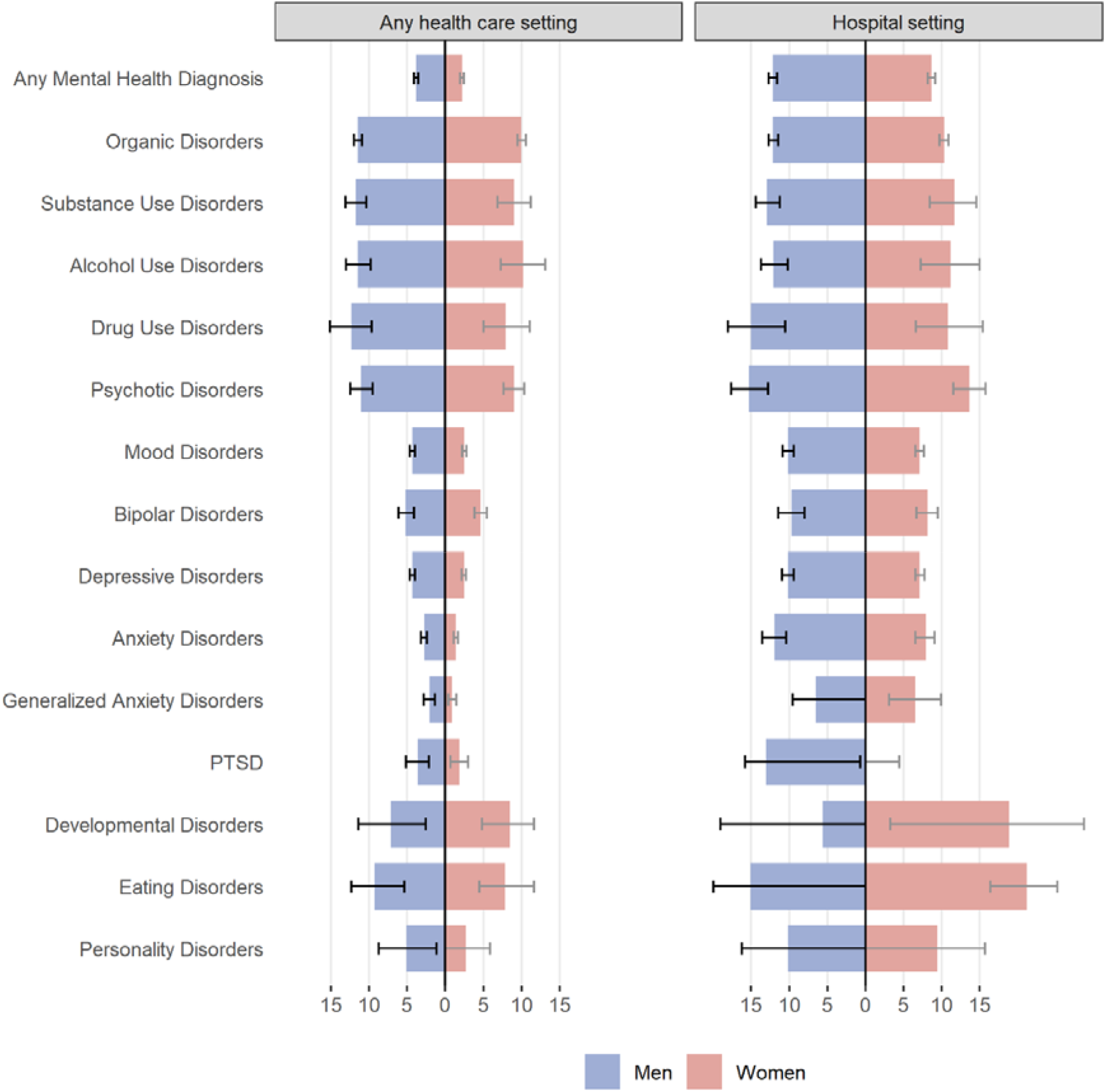
Excess life years lost among people who received mental health diagnoses compared with people of the same sex without mental health diagnoses. Data are stratified by sex and health care setting. Left panel shows data of people diagnosed in any health care setting, such as outpatient or hospital. Right panel shows data from people diagnosed in hospitals. Error bars represent 95% confidence intervals. Confidence limits with negative values are truncated at 0 in the figure. PTSD: Post-traumatic stress disorders

When considering diagnoses from hospital settings only, LYL associated with mental health diagnoses increased to 12.15 years (95% CI 11.59–12.70) among men and 8.67 (95% CI 8.20–9.13) among women (Figure 1, right panel, appendix Table S3). Women hospitalized with an eating disorder (21.17 LYL, 95% CI 16.38–25.18) and men hospitalized with a psychotic disorder (15.29 LYL, 95% CI 12.79–17.61) showed the largest mortality gap.

Most LYL were attributable to natural causes (Tables 3): in men, 3.42 (95% CI 3.17–3.70) LYL associated with any mental health diagnosis were from natural causes, 0.45 (95% CI 0.32–0.59) from unnatural causes, and -0.04 (95% CI -0.11–0.03) from unknown causes; in women, 1.94 (95% CI 1.73–2.15) LYL were from natural causes, 0.22 (95% CI 0.15–0.28) were from unnatural causes, and 0.03 (95% CI -0.02– 0.09) were from unknown causes. Among men, a considerable excess mortality burden was attributable to unnatural causes with alcohol use disorders (LYL 2.49, 95% CI 1.41–3.60), drug use disorders (LYL 2.05, 95% CI 0.58–3.77), and bipolar disorders (LYL 1.52, 95% CI 0.90–2.17). Among women with drug use disorder 1.85 (95% CI 0.34–3.69) LYL were attributable to unnatural causes.

**Table 3:**
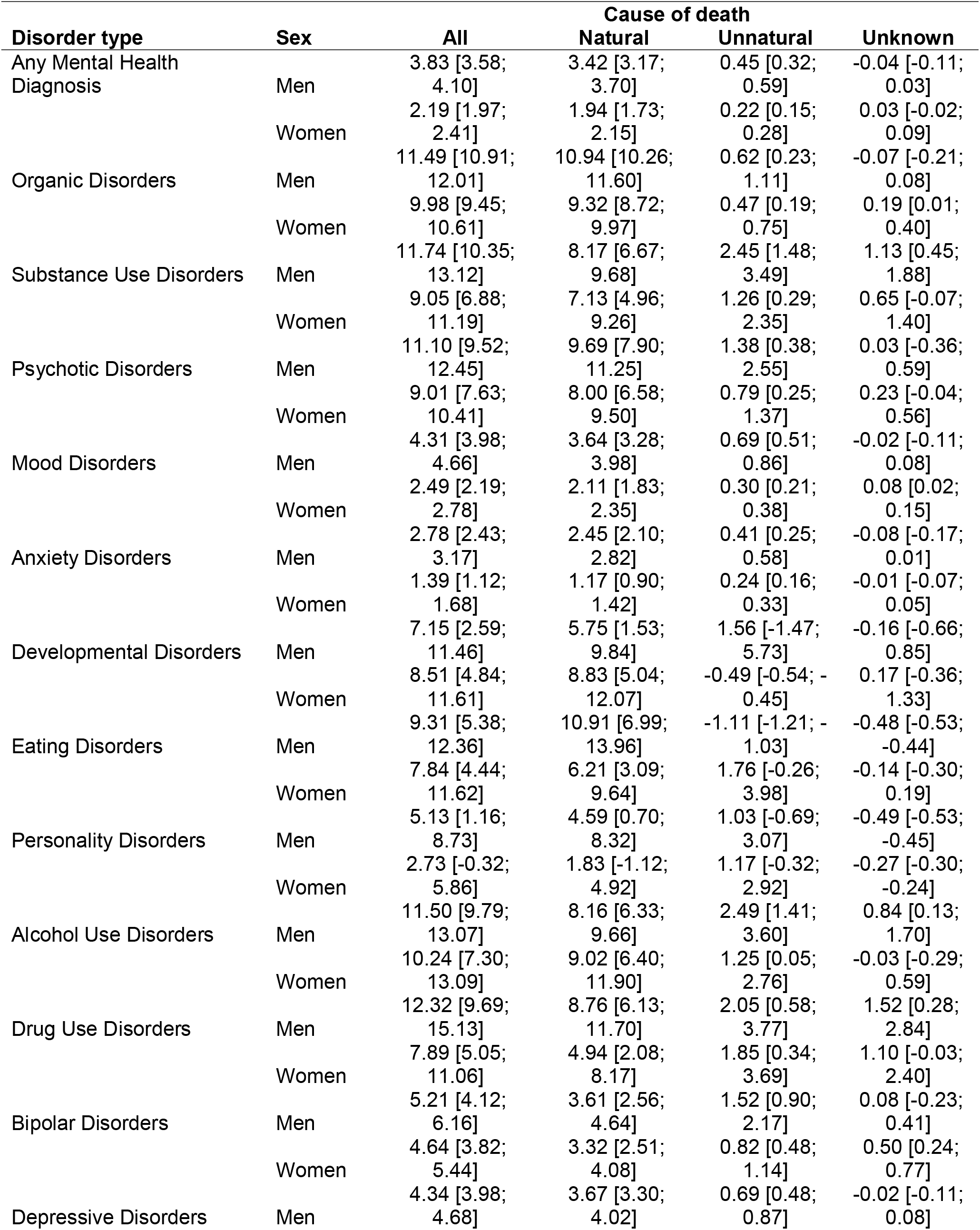
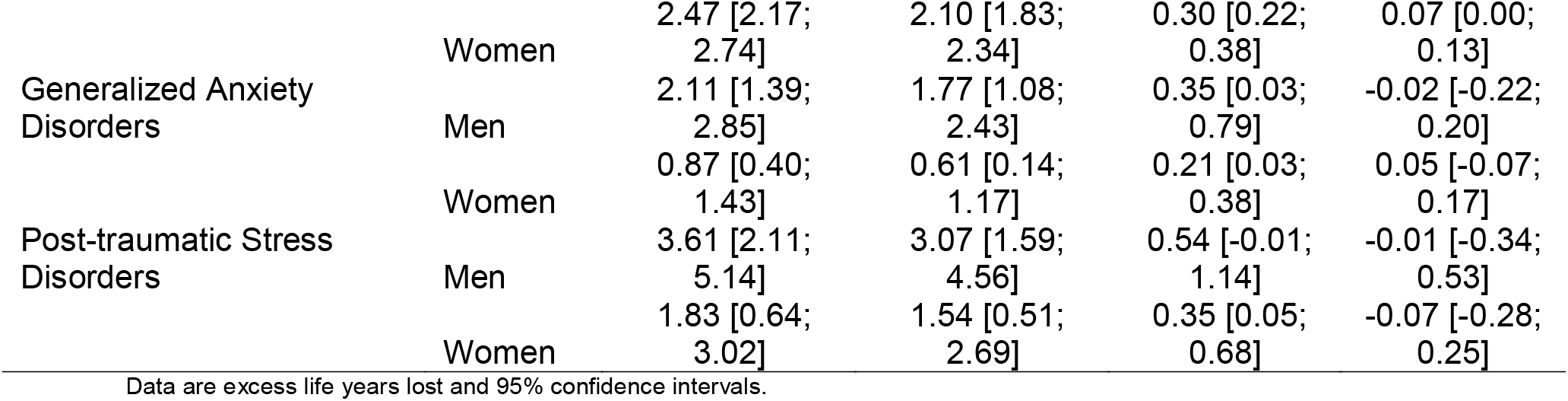
Excess life years lost associated with mental health diagnoses by sex and cause of death

Use of psychiatric medication was associated with 4.05 LYL (95% CI 3.79–4.12) among men and 2.26 LYL (95% CI 2.05–2.48) among women (appendix, Table S4). When limiting exposures to beneficiaries with only one mental health diagnosis, LYL increased to 5.61 years (95% CI 5.17–6.10) among men and 4.41 (95% CI 3.96–4.84) among women (appendix, Table S5). Including deaths occurring within 1 year after an individual’s insurance plan ended did not markedly change results (appendix, Table S6).

Figure 2 shows adjusted HR comparing all-cause mortality of people with and without organic, substance use, psychotic, mood, anxiety, developmental, personality or any mental disorder. The mortality rate was 65% higher among people diagnosed with mental disorders than those without mental health diagnoses (HR 1.65, 95% CI 1.61–1.69). In models adjusted for age, sex, and calendar year (model 1), the greatest increase in mortality risk was among people diagnosed with organic mental disorders (HR 6.01 [95% CI 5.74–6.29]), followed by those diagnosed with psychotic (3.12 [2.83–3.43]), substance use (2.93 [2.64–3.25]), developmental (1.67 [1.31–2.12]), personality (1.65 [1.27–2.15]), mood (1.59 [1.55–1.65]), and anxiety (1.24 [1.20–1.27]) disorders. After further adjusting for psychiatric comorbidity (model 2), associations were attenuated. For example, the HR for psychotic disorders decreased to 1.48 (95% CI 1.34–1.64) and anxiety disorders were no longer associated with all-cause mortality (HR 1.03, 95% CI 0.99–1.06). Excess mortality associated with any mental health diagnosis decreased with follow-up time: from an adjusted HR of 1.96 (95% CI 1.82–2.10) in year 1 of follow-up to an adjusted HR of 1.60 (95% CI 1.56–1.65) beyond the second year of follow-up (appendix, Table S7). Mental health diagnoses of most types were associated with a higher risk of natural and unnatural deaths (appendix, Table S8). Mental health diagnoses during hospitalisations were associated with a 279% increase in mortality risk (HR 3.79, 95% CI 3.66–3.93) (appendix, Table S9). Psychiatric medications were associated with a 77% increase in mortality risk (HR 1.77, 95% CI 1.73–1.81) (appendix, Table S10).

**Figure 2:**
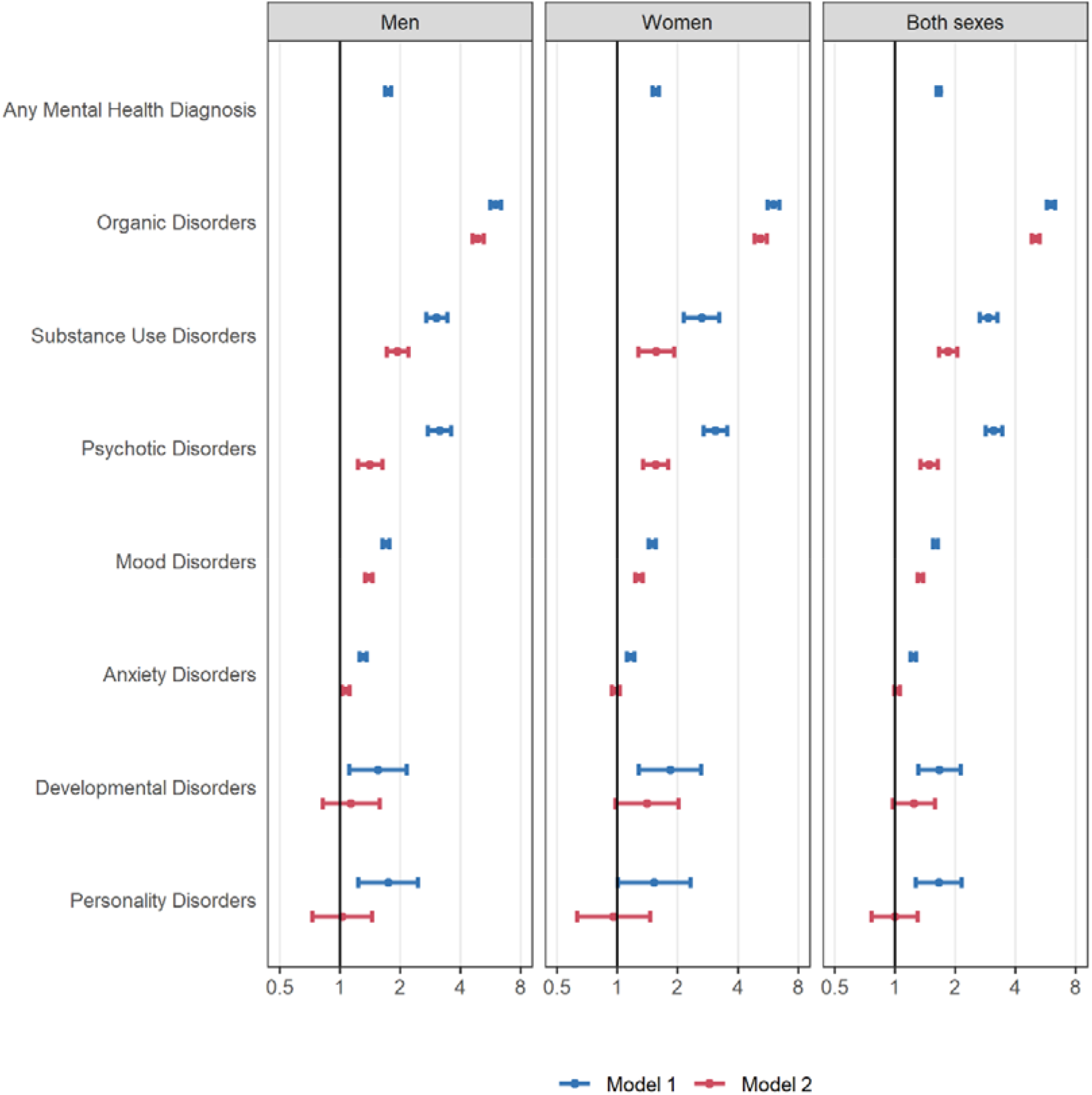
Adjusted hazard ratios comparing all-cause mortality among people with and without mental health diagnoses. We adjusted all models for baseline age, and time-updated calendar year. We adjusted models combining both sexes for sex. Model 1 does not adjust for psychiatric comorbidity; Model 2 adjusts for psychiatric comorbidity. The x-axes are on a logarithmic scale.

## Discussion

In this cohort of South African medical insurance beneficiaries, 22% of men and 31% of women received mental health diagnoses. On average, the remaining life expectancy after a mental health diagnosis was 3.8 years shorter for men and 2.2 years shorter for women compared to people without mental health diagnoses. The difference in life expectancy varied by sex and disorder. Eating disorders, developmental disorders, psychotic disorders, substance use disorders, and organic mental disorders were strongly associated with LYL, especially among men. In contrast, women with anxiety or depression had lower excess mortality. Most excess deaths among people with mental illness were attributable to natural causes.

To our knowledge, this study presents the most comprehensive analysis of excess mortality among people with mental illness in Africa. Earlier studies of excess mortality mainly included people with severe mental illness managed in tertiary care hospitals.^6^ Our study included people with a broad spectrum of disorders managed at all levels of care. We ascertained mortality by linking data from a medical insurance scheme to the South African civil registration system. According to a validation study, the mortality linkage should have identified about 95% of adult deaths.^18^ The mortality data included cause of death information, allowing us to disaggregate excess mortality estimates into deaths attributable to natural and unnatural causes. Finally, we used international diagnostic criteria for the full spectrum of mental disorders to evaluate excess mortality among people with various mental health conditions while adjusting for psychiatric comorbidity.

Although the Global Burden of Disease (GBD) study recognizes mental disorders as leading causes of disease burden, it likely underestimates their true burden.^1,19^ Most mental disorders do not directly lead to death but increase the risk of suicide, accidents, and physical illness, indirectly contributing to premature mortality.^3–6^ In the GBD framework, mortality indirectly attributable to mental disorders is attributed to the cause directly leading to death as recorded on death certificates.^1,20^ For example, suicide resulting from depression will be coded as injury and no mortality burden will be attributed to depression.^1,20^ As a result, the GBD’s disease burden estimates may not reflect the mortality attributable to mental disorders.^1,19^

We calculated an alternative metric, LYL, to quantify excess mortality among people with mental illness. LYL is a descriptive measure of excess mortality reflecting the direct and indirect causal contributions of mental disorders to premature mortality, but also the effect of non-causal confounding factors, for example, precarious living conditions or physical comorbidities influencing both the risk of a mental disorder and mortality. The metric quantifies the average difference in life expectancy after diagnoses of health conditions compared with people of the same age and sex without such diagnoses. LYL are useful for identifying vulnerable populations at high risk of premature mortality who might benefit from targeted interventions to reduce excess mortality. However, LYL must not be interpreted as reflecting the causal effect of mental disorders on mortality.

Although most mental disorders were associated with substantial excess mortality, our estimates were generally lower than those reported in previous studies. A meta-analysis of studies primarily from high-income countries estimated that people diagnosed with a mental disorder died about ten years prematurely.^21^ The overrepresentation of people with severe mental illness in earlier work might partly explain the difference between estimates from earlier studies and our work. Many studies included in the meta-analysis relied on medical records from tertiary care settings, including a large proportion of people with psychotic disorders and other severe mental health conditions associated with a high risk of premature mortality. In contrast, our study included a more representative sample of people with mental illness. We identified most mental health diagnoses based on medical records from outpatient settings, with mental health conditions such as depression and anxiety disorders accounting for most diagnoses. We included people with psychotic disorders and those hospitalized with mental disorders in our study, but persons with severe mental illness were not overrepresented.

People diagnosed with mental illness were at higher risk of death from natural and unnatural causes. Yet, almost 90% of LYL associated with mental disorders were attributable to death from natural causes. Previous studies support this finding. For instance, a register-based cohort study from Denmark attributed 70% (men) and 80% (women) of excess mortality burden to natural causes.^15^ A community-based cohort study from rural Ethiopia attributed 75% of deaths among people with severe mental illness to natural causes.^22^ Higher rates of physical illnesses among people with mental illness and poorer health care contribute to premature death from natural causes among people with mental illness.^3,4,7^ Experts recommend interventions for reducing the burden of physical illness among people with mental health conditions to close the mortality gap.^3^ Interventions can target lifestyle and behaviours to prevent physical illness or early detection and appropriate treatment of common physical comorbidities through screening or collaborative or integrated mental and physical health care models.^3^ Furthermore, interventions aiming to reduce stigma towards mental illness among health care providers should be considered.^23^

For some disorders, unnatural causes of death accounted for considerable excess mortality. For example, among men with alcohol use disorder, drug use disorder, or bipolar disorder, unnatural causes of death accounted for about 20–35% of total LYL. Suicide prevention, substance use treatment, and harm reduction programmes are needed to reduce excess mortality from unnatural causes.^3,24^

Men with mental illness had higher excess mortality than women with mental illness. For alcohol and bipolar disorders, sex disparities in excess mortality are at least partly attributable to higher mortality from unnatural causes among men than women. For other disorders, lower health care utilisation rates among men compared with women,^25^ arising from the gendered nature of health services creating health care barriers for men,^26,27^ harmful masculine norms,^28^ self-stigmatizing beliefs,^29^ or differences in coping strategies^30^ may contribute to higher excess mortality for men than women. Further work is needed to design, implement, and evaluate strategies addressing men’s physical and mental health care needs.

Our study has several limitations. First, we assessed beneficiaries’ mental health status based on ICD-10 diagnoses from reimbursement claims; thus, we missed beneficiaries with undiagnosed mental disorders who may be less severely ill than those diagnosed. Furthermore, mental health diagnoses might be affected by diagnostic or administrative errors,^31^ although diagnoses from administrative data generally have a high positive predictive value for research diagnoses.^31^ Sensitivity analysis of people with only one, possibly erroneous, mental health diagnosis showed high excess mortality, suggesting this population might constitute people with untreated conditions rather than false positive cases. Second, we did not consider remissions. This could have led to an underestimation of the excess mortality, particularly for depression or anxiety. Third, our study only included data from a private-sector medical insurance scheme. Thus, our findings do not necessarily apply to people accessing the public health care sector. People using public health services generally have lower socioeconomic status. They may be at higher risk of experiencing poor mental health outcomes than people who access private services.^32^ Access to mental health care in South Africa’s public sector is limited and mental disorders often remain untreated.^33^

In conclusion, our study demonstrates a considerable burden of premature death among people with mental illness. People diagnosed with eating, developmental, psychotic, substance use, and organic mental disorders—especially men and those hospitalized with these conditions—are at the highest risk of premature death. These findings support implementing interventions for prevention, early detection, and appropriate treatment of physical comorbidities among people with mental illness. In addition, suicide prevention and substance use treatment programmes are needed to reduce excess mortality from unnatural causes among men with bipolar and substance use disorders.

## Data Availability

Data were obtained from the International epidemiology Databases to Evaluate AIDS-Southern Africa (IeDEA-SA). Data cannot be made available online because of legal and ethical restrictions. To request data, readers may contact IeDEA-SA for consideration by filling out the online form available at https://www.iedea-sa.org/contact-us/. Statistical code is available under https://github.com/IeDEA-SA/LYL

## Authors and contributors

YR and AH conceived the study. ME and AH obtained funding for the study. YR, AW and AH wrote the first draft of the study protocol, which was revised by all authors. YR and AW did the analyses. CW, LJ, MS and CD advised on statistical analyses. AW, YR, and AH wrote the first draft of the paper. All other authors contributed to the interpretation of results and critically revised the manuscript for intellectual content. YR, AW and AH had access to and verified the underlying data reported in the manuscript. All authors have approved the final version of the manuscript for submission for publication. Kristin Marie Bivens, PhD from the Institute of Social and Preventive Medicine at the University of Bern, edited the manuscript.

## Data sharing

Data were obtained from the International epidemiology Databases to Evaluate AIDS–Southern Africa (IeDEA-SA). Data cannot be made available online because of legal and ethical restrictions. To request data, readers may contact IeDEA-SA for consideration by filling out the online form available at https://www.iedea-sa.org/contact-us/. Statistical code is available under https://github.com/IeDEA-SA/LYL

## Ethical considerations

The Human Research Ethics Committee of the University of Cape Town, South Africa and the Cantonal Ethics Committee, Bern, Switzerland, granted ethical permission for the analysis. Beneficiaries of the medical scheme or their guardians consented to the use of their data in research.

## Additional Contributions

Calculations were performed on UBELIX (http://www.id.unibe.ch/hpc), the high-performance computing cluster at the University of Bern.

