## Appendix for "Life-years lost associated with mental illness: a cohort study of beneficiaries of a South African medical insurance scheme"

**Figure S1:** Flow diagram showing selection of eligible individuals for analyses.

**Figure S2:** Psychiatric comorbidity among beneficiaries with a mental health diagnosis.

**Table S1:** Number and proportion of people who received mental health diagnoses in hospital settings during follow-up by sex.

**Table S2**: Number and proportion of people who received psychiatric medication during follow-up by sex.

**Table S3**: Excess life years lost associated with mental health diagnoses from hospital settings by sex and cause of death.

**Table S4:** Excess life years lost associated with psychiatric medication by sex and cause of death.

**Table S5**: Excess life years lost associated with mental health diagnoses by sex and cause of death. The exposed groups include only individuals with exactly one diagnosis of the given type.

**Table S6**: Excess life years lost associated with mental health diagnoses by sex and cause of death. People who died within one year of the end of their insurance coverage are not censored at the end of the coverage.

**Table S7**: Hazard ratios per follow-up time interval comparing all-cause mortality among individuals with mental health diagnoses to those without.

**Table S8**: Hazard ratios (HR) comparing mortality (all-cause, natural deaths, unnatural deaths) among individuals with mental health diagnoses to those without unadjusted and adjusted for psychiatric comorbidity.

**Table S9**: Hazard ratios (HR) comparing mortality (all-cause and natural and unnatural deaths) among individuals with mental health diagnoses from hospital settings with those without unadjusted and adjusted for psychiatric comorbidity.

**Table S10**: Hazard ratios (HR) comparing mortality (all-cause, natural deaths, unnatural deaths) among individuals prescribed psychiatric medication to those who were not prescribed psychiatric medication unadjusted and adjusted for psychiatric comorbidity.

**Figure S1:** Flow diagram showing selection of eligible individuals for analyses.

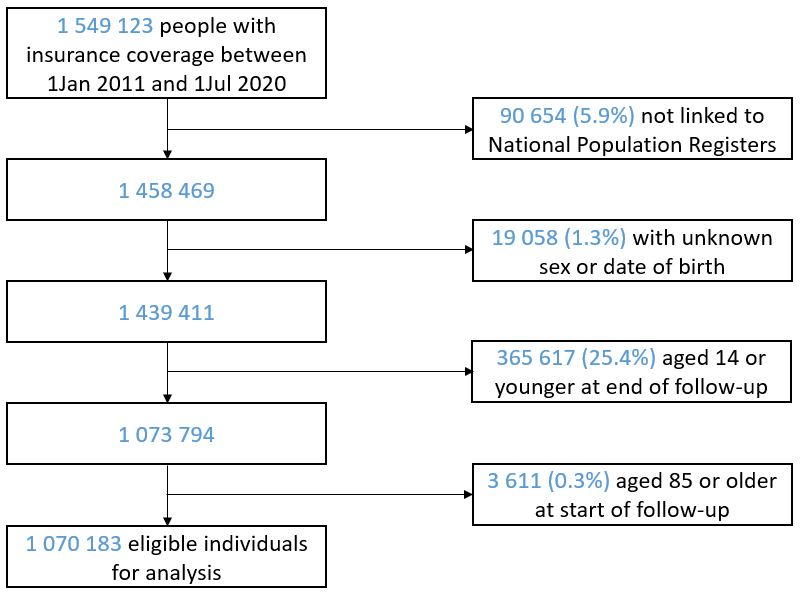

**Figure S2:** Psychiatric comorbidity among beneficiaries with a mental health diagnosis.

Percentage of beneficiaries with diagnoses shown on the x-axis among those with diagnoses shown on the y-axis. Darker colours represent higher values. Org=organic mental disorders, SU=substance use disorders, psy=psychotic disorders, mood=mood disorders, anx=anxiety disorders, dev=developmental disorders, pers=personality disorders.

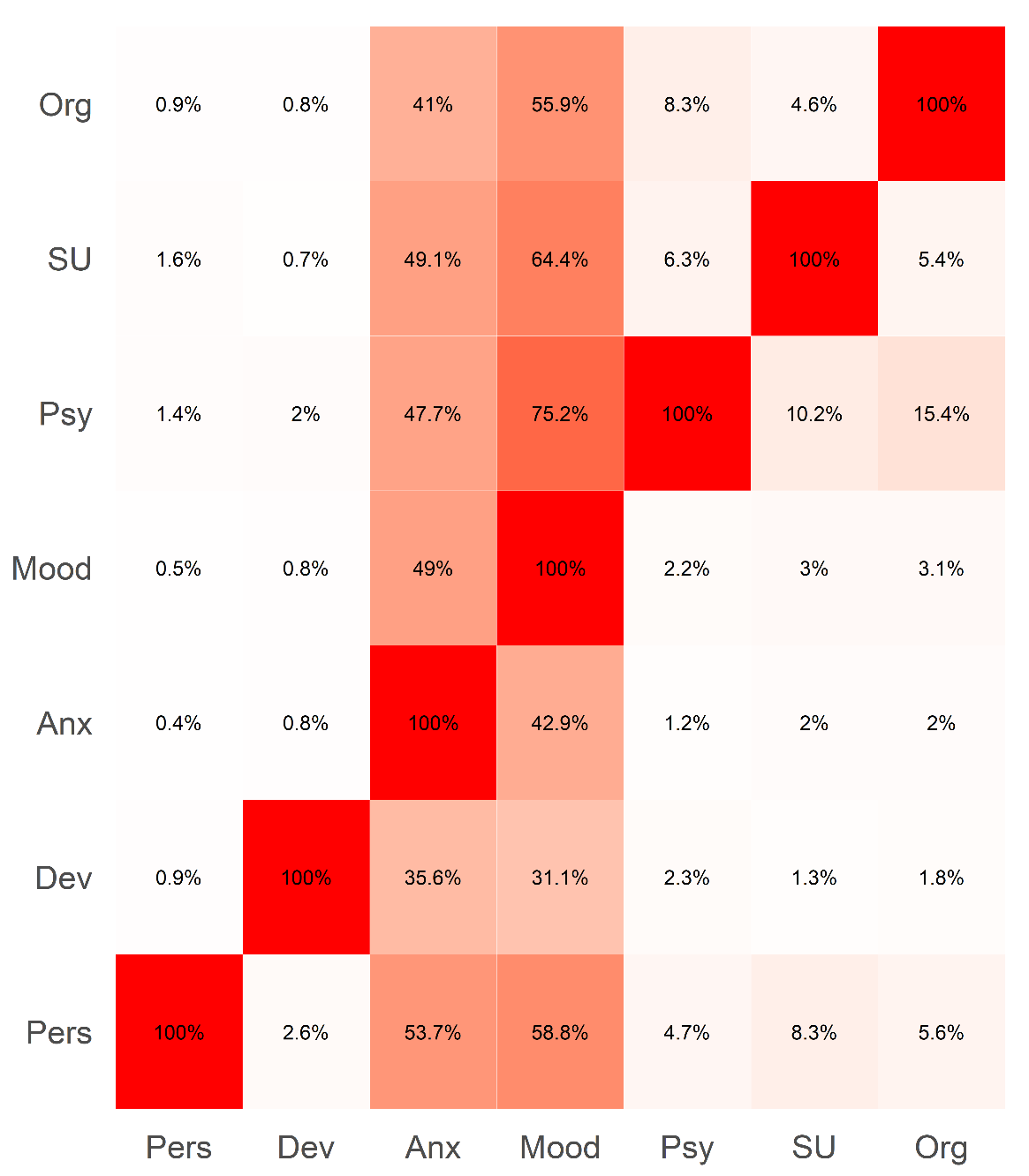

**Table S1:** Number and proportion of people who received mental health diagnoses in hospital settings during follow-up by sex.

|  | **Beneficiaries, No. (%)** | | |
| --- | --- | --- | --- |
|  | **Men** | **Women** | **Total** |
|  | N=517 305 | N=552 878 | N=1 070 183 |
| Any Mental Health Diagnosis (F00-F99) | 17 412 (3.4) | 28 263 (5.1) | 45 675 (4.3) |
| Organic Mental Disorders (F00-F09) | 1 951 (0.4) | 2 131 (0.4) | 4 082 (0.4) |
| Substance Use Disorders (F10-F17, F19) | 4 093 (0.8) | 1 216 (0.2) | 5 309 (0.5) |
| Alcohol Use Disorders (F10) | 2 145 (0.4) | 596 (0.1) | 2 741 (0.3) |
| Drug Use Disorders (F11-F17, F19) | 2 236 (0.4) | 687 (0.1) | 2 923 (0.3) |
| Psychotic Disorders (F20-F29) | 939 (0.2) | 863 (0.2) | 1 802 (0.2) |
| Mood Disorders (F30-F39) | 12 796 (2.5) | 24 752 (4.5) | 37 548 (3.5) |
| Bipolar Disorders (F31) | 2 351 (0.5) | 4 355 (0.8) | 6 706 (0.6) |
| Depression (F32-F33, F34.1) | 11 590 (2.2) | 22 803 (4.1) | 34 393 (3.2) |
| Anxiety Disorders (F40-F48) | 2 123 (0.4) | 5 058 (0.9) | 7 181 (0.7) |
| Generalized Anxiety Disorders (F41.1) | 373 (0.1) | 762 (0.1) | 1 135 (0.1) |
| Post-traumatic Stress Disorders (F43.1) | 421 (0.1) | 964 (0.2) | 1 385 (0.1) |
| Developmental Disorders (F80-F89) | 58 (0.0) | 44 (0.0) | 102 (0.0) |
| Eating Disorders (F51) | 20 (0.0) | 91 (0.0) | 111 (0.0) |
| Personality Disorders (F60-F69) | 105 (0.0) | 157 (0.0) | 262 (0.0) |

International Classification of Diseases, tenth revision code range of each diagnostic category is shown in parenthesis.

**Table S2**: Number and proportion of people who received psychiatric medication during follow-up by sex.

|  | **Beneficiaries, No. (%)** | | |
| --- | --- | --- | --- |
|  | **Men** | **Women** | **Total** |
|  | N=517 305 | N=552 878 | N=1 070 183 |
| Any Psychiatric Medication | 148 221 (28.7) | 224 096 (40.5) | 372 317 (34.8) |
| Substance Use Medication | 3 262 (0.6) | 2 281 (0.4) | 5 543 (0.5) |
| Antidepressant | 89 882 (17.4) | 148 475 (26.9) | 238 357 (22.3) |
| Anxiolytic | 90 114 (17.4) | 146 137 (26.4) | 236 251 (22.1) |
| Antipsychotic | 37 002 (7.2) | 64 707 (11.7) | 10 1709 (9.5) |

**Table S3**: Excess life years lost associated with mental health diagnoses from hospital settings by sex and cause of death.

|  | | **Cause of death** | | | |
| --- | --- | --- | --- | --- | --- |
| **Disorder type** | **Sex** | **All** | **Natural** | **Unnatural** | **Unknown** |
| Any Mental Health Diagnosis | Men | 12.15 [11.59; 12.70] | 10.40 [9.75; 10.97] | 1.68 [1.27; 2.15] | 0.07 [-0.10; 0.28] |
|  | Women | 8.67 [ 8.20; 9.13] | 7.71 [7.25; 8.18] | 0.70 [0.49; 0.90] | 0.26 [0.11; 0.40] |
| Organic Disorders | Men | 12.12 [11.46; 12.70] | 11.86 [11.10; 12.52] | 0.29 [-0.09; 0.78] | -0.02 [-0.19; 0.19] |
|  | Women | 10.32 [ 9.71; 10.95] | 9.91 [9.27; 10.62] | 0.24 [0.03; 0.44] | 0.17 [0.00; 0.36] |
| Substance Use Disorders | Men | 12.94 [11.24; 14.38] | 9.00 [7.09; 10.62] | 2.56 [1.36; 3.68] | 1.38 [0.50; 2.37] |
|  | Women | 11.67 [ 8.47; 14.60] | 8.59 [5.44; 11.80] | 2.06 [0.47; 3.74] | 1.02 [-0.09; 2.22] |
| Psychotic Disorders | Men | 15.29 [12.79; 17.61] | 13.96 [10.99; 16.55] | 1.82 [0.11; 3.87] | -0.50 [-0.54; -0.44] |
|  | Women | 13.69 [11.54; 15.75] | 12.29 [9.93; 14.91] | 1.52 [0.39; 2.62] | -0.12 [-0.26; 0.21] |
| Mood Disorders | Men | 10.17 [ 9.42; 10.90] | 8.36 [7.62; 9.04] | 1.77 [1.27; 2.31] | 0.04 [-0.17; 0.27] |
|  | Women | 7.13 [ 6.54; 7.69] | 6.10 [5.54; 6.66] | 0.76 [0.51; 0.97] | 0.27 [0.10; 0.43] |
| Anxiety Disorders | Men | 11.96 [10.37; 13.58] | 9.86 [8.28; 11.49] | 2.04 [0.97; 3.41] | 0.06 [-0.37; 0.58] |
|  | Women | 7.97 [ 6.53; 9.08] | 6.63 [5.22; 7.88] | 1.04 [0.50; 1.69] | 0.29 [-0.11; 0.76] |
| Developmental Disorders | Men | 5.63 [-10.83; 19.03] | 7.83 [-8.61; 21.21] | -1.66 [-1.89; -1.40] | -0.54 [-0.60; -0.48] |
|  | Women | 18.84 [ 3.24; 28.71] | 9.62 [-4.49; 24.80] | -0.37 [-0.47; -0.28] | 9.59 [-0.30; 26.08] |
| Eating Disorders | Men | 15.14 [-10.88; 19.97] | 16.81 [-8.94; 21.72] | -1.18 [-1.54; -0.84] | -0.49 [-0.56; -0.42] |
|  | Women | 21.17 [16.38; 25.18] | 11.07 [2.81; 17.27] | 8.91 [-0.40; 18.71] | 1.19 [-0.32; 4.82] |
| Personality Disorders | Men | 10.19 [-2.39; 16.26] | 6.41 [-4.36; 12.84] | 4.28 [-1.17; 11.07] | -0.50 [-0.55; -0.45] |
|  | Women | 9.41 [-0.34; 15.68] | 6.45 [-2.63; 12.56] | 3.24 [-0.35; 8.67] | -0.28 [-0.31; -0.25] |
| Alcohol Use Disorders | Men | 12.06 [10.20; 13.73] | 8.74 [6.79; 10.45] | 2.25 [1.09; 3.42] | 1.07 [0.18; 2.17] |
|  | Women | 11.22 [ 7.29; 15.03] | 8.69 [4.45; 12.82] | 2.32 [0.29; 4.68] | 0.21 [-0.29; 1.41] |
| Drug Use Disorders | Men | 15.01 [10.53; 18.04] | 11.34 [7.00; 14.38] | 2.19 [0.42; 4.36] | 1.48 [0.10; 3.07] |
|  | Women | 10.83 [ 6.63; 15.44] | 7.10 [2.97; 12.05] | 1.92 [0.11; 4.26] | 1.80 [-0.27; 4.11] |
| Bipolar Disorders | Men | 9.69 [ 8.00; 11.45] | 6.02 [4.14; 7.96] | 3.04 [1.62; 4.65] | 0.63 [-0.19; 1.54] |
|  | Women | 8.17 [ 6.68; 9.51] | 5.80 [4.33; 7.16] | 1.64 [0.90; 2.33] | 0.74 [0.18; 1.36] |
| Depressive Disorders | Men | 10.21 [ 9.42; 10.94] | 8.52 [7.67; 9.22] | 1.71 [1.20; 2.26] | -0.02 [-0.22; 0.20] |
|  | Women | 7.15 [ 6.57; 7.75] | 6.17 [5.59; 6.77] | 0.70 [0.45; 0.94] | 0.27 [0.10; 0.45] |
| Generalized Anxiety Disorders | Men | 6.55 [-4.17; 9.56] | 7.19 [-3.33; 10.34] | -0.14 [-1.23; 2.11] | -0.50 [-0.54; -0.46] |
|  | Women | 6.58 [ 3.12; 9.94] | 5.45 [1.77; 8.65] | 0.70 [-0.31; 2.26] | 0.43 [-0.29; 2.53] |
| Post-traumatic Stress Disorders | Men | 13.08 [ 0.70; 15.84] | 7.36 [-4.82; 9.96] | 4.32 [1.14; 7.70] | 1.40 [-0.50; 3.59] |
|  | Women | 0.05 [-3.64; 4.45] | -2.17 [-5.13; 1.67] | 2.51 [0.51; 5.06] | -0.28 [-0.31; -0.25] |

**Table S4:** Excess life years lost associated with psychiatric medication by sex and cause of death.

|  | | **Cause of death** | | | |
| --- | --- | --- | --- | --- | --- |
| **Medication type** | **Sex** | **All** | **Natural** | **Unnatural** | **Unknown** |
| Any | Men | 4.05 [3.79; 4.27] | 3.90 [3.66; 4.12] | 0.22 [ 0.13; 0.32] | -0.07 [-0.14; 0.00] |
|  | Women | 2.26 [2.05; 2.48] | 2.13 [1.94; 2.34] | 0.10 [ 0.04; 0.15] | 0.03 [-0.02; 0.07] |
| Antidepressant | Men | 4.49 [4.22; 4.77] | 4.20 [3.92; 4.46] | 0.36 [ 0.25; 0.49] | -0.07 [-0.14; 0.01] |
|  | Women | 2.50 [2.27; 2.74] | 2.32 [2.11; 2.54] | 0.14 [ 0.08; 0.20] | 0.04 [-0.02; 0.09] |
| Antipsychotic | Men | 7.16 [6.76; 7.53] | 6.77 [6.38; 7.13] | 0.35 [ 0.16; 0.55] | 0.03 [-0.09; 0.14] |
|  | Women | 4.53 [4.21; 4.88] | 4.18 [3.89; 4.53] | 0.25 [ 0.15; 0.36] | 0.09 [ 0.01; 0.17] |
| Anxiolytic | Men | 3.85 [3.57; 4.10] | 3.73 [3.47; 4.00] | 0.19 [ 0.07; 0.30] | -0.07 [-0.16; 0.01] |
|  | Women | 2.20 [1.96; 2.44] | 2.05 [1.83; 2.28] | 0.14 [ 0.07; 0.20] | 0.02 [-0.04; 0.07] |
| Substance Use | Men | 3.34 [1.42; 5.03] | 3.42 [1.64; 5.20] | 0.07 [-0.54; 0.83] | -0.15 [-0.47; 0.23] |
|  | Women | 4.36 [2.41; 6.33] | 4.15 [2.19; 6.02] | 0.13 [-0.27; 0.64] | 0.09 [-0.26; 0.68] |

**Table S5**: Excess life years lost associated with mental health diagnoses by sex and cause of death. The exposed groups include only individuals with exactly one diagnosis of the given type.

|  | | **Cause of death** | | | |
| --- | --- | --- | --- | --- | --- |
| **Disorder type** | **Sex** | **All** | **Natural** | **Unnatural** | **Unknown** |
| Any Mental Health Diagnosis | Men | 5.61 [ 5.17; 6.10] | 5.09 [ 4.63; 5.55] | 0.44 [ 0.21; 0.69] | 0.08 [-0.06; 0.24] |
|  | Women | 4.41 [ 3.96; 4.84] | 4.04 [ 3.60; 4.49] | 0.36 [ 0.21; 0.51] | 0.02 [-0.09; 0.12] |
| Organic Disorders | Men | 13.32 [12.50; 14.19] | 12.29 [11.26; 13.30] | 1.08 [ 0.33; 1.94] | -0.06 [-0.27; 0.22] |
|  | Women | 11.70 [10.84; 12.64] | 11.04 [10.14; 12.00] | 0.46 [ 0.07; 0.88] | 0.20 [-0.04; 0.49] |
| Substance Use Disorders | Men | 12.41 [10.23; 14.90] | 9.99 [ 7.38; 12.42] | 2.04 [ 0.46; 3.83] | 0.38 [-0.43; 1.46] |
|  | Women | 8.85 [ 5.60; 12.23] | 7.71 [ 4.47; 11.24] | 0.62 [-0.33; 1.97] | 0.52 [-0.28; 1.60] |
| Psychotic Disorders | Men | 12.11 [ 9.84; 14.39] | 11.15 [ 8.86; 13.40] | 0.95 [-0.69; 2.83] | 0.00 [-0.48; 1.03] |
|  | Women | 10.69 [ 7.94; 13.07] | 9.84 [ 6.99; 12.47] | 0.53 [-0.27; 1.58] | 0.31 [-0.25; 1.03] |
| Mood Disorders | Men | 6.53 [ 5.83; 7.14] | 5.74 [ 5.05; 6.33] | 0.78 [ 0.44; 1.10] | 0.01 [-0.15; 0.19] |
|  | Women | 4.66 [ 4.01; 5.25] | 4.16 [ 3.56; 4.75] | 0.37 [ 0.18; 0.56] | 0.13 [-0.03; 0.29] |
| Anxiety Disorders | Men | 4.24 [ 3.76; 4.79] | 3.64 [ 3.14; 4.13] | 0.56 [ 0.31; 0.84] | 0.05 [-0.10; 0.20] |
|  | Women | 2.91 [ 2.43; 3.33] | 2.58 [ 2.11; 3.00] | 0.41 [ 0.26; 0.57] | -0.08 [-0.16; 0.01] |
| Developmental Disorders | Men | 3.26 [-3.27; 9.52] | 4.74 [-1.42; 11.39] | -0.89 [-1.90; 1.42] | -0.59 [-0.64; -0.54] |
|  | Women | 6.53 [ 1.17; 11.25] | 6.50 [ 1.27; 11.52] | -0.42 [-0.47; -0.38] | 0.46 [-0.34; 2.31] |
| Eating Disorders | Men | 10.64 [ 6.05; 13.64] | 12.29 [ 7.69; 15.32] | -1.16 [-1.26; -1.06] | -0.49 [-0.53; -0.45] |
|  | Women | 7.46 [ 2.96; 11.39] | 6.40 [ 2.24; 10.21] | 1.34 [-0.33; 4.29] | -0.28 [-0.31; -0.24] |
| Personality Disorders | Men | 5.01 [ 0.59; 9.28] | 4.58 [ 0.04; 8.41] | 0.92 [-0.88; 3.28] | -0.49 [-0.53; -0.45] |
|  | Women | 3.32 [-0.44; 7.35] | 1.78 [-1.46; 5.60] | 1.82 [-0.32; 4.36] | -0.27 [-0.30; -0.24] |
| Alcohol Use Disorders | Men | 12.62 [ 9.21; 15.42] | 8.50 [ 5.33; 11.61] | 3.15 [ 1.08; 5.37] | 0.98 [-0.18; 2.57] |
|  | Women | 9.27 [ 4.33; 14.37] | 7.30 [ 2.72; 12.33] | 2.24 [-0.33; 5.95] | -0.27 [-0.30; -0.24] |
| Drug Use Disorders | Men | 14.46 [10.53; 17.03] | 12.84 [ 8.32; 15.80] | 2.15 [-0.12; 5.26] | -0.53 [-0.58; -0.06] |
|  | Women | 7.68 [ 2.97; 12.04] | 6.43 [ 1.78; 11.29] | 0.39 [-0.34; 1.79] | 0.86 [-0.28; 2.45] |
| Bipolar Disorders | Men | 4.79 [ 2.69; 6.74] | 3.37 [ 1.12; 5.39] | 1.36 [ 0.12; 2.67] | 0.05 [-0.45; 0.86] |
|  | Women | 7.04 [ 5.46; 8.64] | 5.47 [ 3.95; 7.12] | 0.81 [ 0.17; 1.55] | 0.77 [ 0.11; 1.42] |
| Depressive Disorders | Men | 6.53 [ 5.85; 7.14] | 5.77 [ 5.07; 6.39] | 0.76 [ 0.43; 1.10] | 0.00 [-0.17; 0.17] |
|  | Women | 4.70 [ 4.07; 5.30] | 4.25 [ 3.67; 4.86] | 0.35 [ 0.16; 0.53] | 0.11 [-0.04; 0.26] |
| Generalized Anxiety Disorders | Men | 2.53 [ 1.43; 3.52] | 2.38 [ 1.35; 3.39] | 0.18 [-0.28; 0.68] | -0.03 [-0.31; 0.30] |
|  | Women | 1.14 [ 0.47; 2.05] | 1.02 [ 0.28; 1.92] | 0.14 [-0.06; 0.36] | -0.01 [-0.16; 0.16] |
| Post-traumatic Stress Disorders | Men | 3.82 [ 1.84; 5.73] | 2.94 [ 1.13; 4.81] | 0.84 [ 0.05; 1.70] | 0.04 [-0.39; 0.53] |
|  | Women | 1.04 [-0.48; 2.60] | 0.93 [-0.43; 2.55] | 0.21 [-0.12; 0.61] | -0.10 [-0.29; 0.32] |

**Table S6**: Excess life years lost associated with mental health diagnoses by sex and cause of death. People who died within one year of the end of their insurance coverage are not censored at the end of the coverage.

|  | | **Cause of death** | | | |
| --- | --- | --- | --- | --- | --- |
| **Disorder type** | **Sex** | **All** | **Natural** | **Unnatural** | **Unknown** |
| Any Mental Health Diagnosis | Men | 3.69 [ 3.41; 3.96] | 3.21 [ 2.94; 3.48] | 0.50 [ 0.35; 0.65] | -0.02 [-0.09; 0.06] |
|  | Women | 2.07 [ 1.87; 2.29] | 1.82 [ 1.61; 2.04] | 0.22 [ 0.15; 0.29] | 0.03 [-0.02; 0.09] |
| Organic Disorders | Men | 11.27 [10.75; 11.90] | 10.72 [10.01; 11.38] | 0.63 [ 0.18; 1.16] | -0.08 [-0.21; 0.09] |
|  | Women | 10.02 [ 9.47; 10.61] | 9.39 [ 8.76; 10.00] | 0.47 [ 0.18; 0.75] | 0.17 [ 0.01; 0.37] |
| Substance Use Disorders | Men | 12.04 [10.59; 13.35] | 7.85 [ 6.44; 9.36] | 3.08 [ 2.04; 4.12] | 1.11 [ 0.37; 1.85] |
|  | Women | 9.63 [ 7.67; 11.55] | 7.50 [ 5.38; 9.64] | 1.34 [ 0.45; 2.32] | 0.79 [ 0.06; 1.62] |
| Psychotic Disorders | Men | 11.39 [ 9.88; 12.80] | 9.79 [ 8.44; 11.36] | 1.60 [ 0.60; 2.88] | 0.00 [-0.39; 0.50] |
|  | Women | 9.35 [ 8.03; 10.63] | 8.29 [ 6.88; 9.65] | 0.81 [ 0.24; 1.47] | 0.25 [-0.04; 0.62] |
| Mood Disorders | Men | 4.15 [ 3.79; 4.50] | 3.39 [ 3.05; 3.73] | 0.76 [ 0.57; 0.96] | 0.00 [-0.09; 0.11] |
|  | Women | 2.33 [ 2.08; 2.62] | 1.94 [ 1.68; 2.22] | 0.31 [ 0.21; 0.40] | 0.08 [ 0.01; 0.15] |
| Anxiety Disorders | Men | 2.59 [ 2.26; 2.96] | 2.23 [ 1.91; 2.59] | 0.43 [ 0.25; 0.59] | -0.07 [-0.15; 0.03] |
|  | Women | 1.31 [ 1.04; 1.57] | 1.08 [ 0.83; 1.35] | 0.24 [ 0.14; 0.34] | -0.02 [-0.08; 0.05] |
| Developmental Disorders | Men | 7.15 [ 2.67; 12.12] | 5.20 [ 1.52; 9.03] | 2.18 [-1.38; 7.02] | -0.23 [-0.66; 0.72] |
|  | Women | 7.86 [ 4.17; 11.19] | 8.28 [ 4.52; 11.40] | -0.58 [-0.63; -0.53] | 0.16 [-0.36; 1.33] |
| Eating Disorders | Men | 8.48 [ 5.44; 11.68] | 10.28 [ 7.22; 13.49] | -1.32 [-1.42; -1.21] | -0.48 [-0.52; -0.44] |
|  | Women | 7.39 [ 4.07; 10.40] | 5.84 [ 2.61; 9.01] | 1.69 [-0.03; 4.21] | -0.14 [-0.31; 0.21] |
| Personality Disorders | Men | 4.76 [ 1.31; 8.12] | 3.59 [ 0.42; 7.11] | 1.29 [-0.58; 3.66] | -0.12 [-0.52; 0.78] |
|  | Women | 3.29 [-0.13; 6.95] | 2.50 [-0.86; 5.99] | 1.07 [-0.36; 2.90] | -0.27 [-0.31; -0.24] |
| Alcohol Use Disorders | Men | 11.80 [10.23; 13.30] | 8.14 [ 6.62; 9.91] | 2.90 [ 1.78; 4.13] | 0.75 [ 0.04; 1.57] |
|  | Women | 10.84 [ 7.94; 13.69] | 9.48 [ 6.44; 12.44] | 1.41 [ 0.15; 2.87] | -0.05 [-0.29; 0.54] |
| Drug Use Disorders | Men | 12.31 [ 9.29; 14.92] | 7.57 [ 4.44; 10.43] | 3.09 [ 1.26; 4.91] | 1.66 [ 0.32; 3.15] |
|  | Women | 8.20 [ 5.39; 11.34] | 5.07 [ 2.46; 8.04] | 1.74 [ 0.29; 3.62] | 1.38 [ 0.11; 2.78] |
| Bipolar Disorders | Men | 5.36 [ 4.48; 6.39] | 3.54 [ 2.60; 4.63] | 1.67 [ 0.96; 2.39] | 0.15 [-0.16; 0.48] |
|  | Women | 4.54 [ 3.69; 5.36] | 3.15 [ 2.43; 3.92] | 0.86 [ 0.55; 1.19] | 0.52 [ 0.25; 0.86] |
| Depressive Disorders | Men | 4.17 [ 3.83; 4.53] | 3.42 [ 3.06; 3.77] | 0.76 [ 0.57; 0.97] | -0.01 [-0.10; 0.11] |
|  | Women | 2.31 [ 2.05; 2.61] | 1.94 [ 1.67; 2.24] | 0.31 [ 0.21; 0.40] | 0.06 [-0.01; 0.13] |
| Generalized Anxiety Disorders | Men | 1.81 [ 1.07; 2.50] | 1.44 [ 0.71; 2.18] | 0.40 [ 0.02; 0.78] | -0.03 [-0.22; 0.18] |
|  | Women | 0.71 [ 0.18; 1.20] | 0.43 [-0.04; 0.92] | 0.22 [ 0.06; 0.41] | 0.06 [-0.07; 0.21] |
| Post-traumatic Stress Disorders | Men | 3.67 [ 2.18; 5.08] | 3.10 [ 1.60; 4.71] | 0.59 [ 0.01; 1.23] | -0.02 [-0.39; 0.41] |
|  | Women | 1.72 [ 0.55; 2.79] | 1.43 [ 0.43; 2.46] | 0.36 [ 0.05; 0.79] | -0.07 [-0.29; 0.26] |

**Table S7**: Hazard ratios per follow-up time interval comparing all-cause mortality among individuals with mental health diagnoses to those without.

|  | **Year of follow-up** | | |
| --- | --- | --- | --- |
| **Disorder type** | **First** | **Second** | **Third and beyond** |
| Any Mental Health Diagnosis | 1.96 [1.82; 2.10] | 1.72 [1.61; 1.84] | 1.60 [1.56; 1.65] |
| Organic Disorders | 8.12 [6.85; 9.63] | 6.31 [5.48; 7.28] | 4.74 [4.49; 5.00] |
| Substance Use Disorders | 3.16 [2.19; 4.57] | 1.98 [1.39; 2.81] | 1.77 [1.58; 1.99] |
| Psychotic Disorders | 1.81 [1.26; 2.61] | 1.58 [1.14; 2.18] | 1.47 [1.32; 1.64] |
| Mood Disorders | 1.53 [1.38; 1.69] | 1.42 [1.29; 1.55] | 1.30 [1.26; 1.35] |
| Anxiety Disorders | 1.11 [0.98; 1.26] | 1.06 [0.96; 1.17] | 1.02 [0.99; 1.06] |
| Developmental Disorders | 1.06 [0.39; 2.86] | 1.23 [0.55; 2.76] | 1.28 [0.98; 1.66] |
| Personality Disorders | 1.55 [0.58; 4.16] | 0.20 [0.03; 1.39] | 1.06 [0.80; 1.40] |

The models were adjusted for sex, baseline age, psychiatric comorbidity, and time-updated calendar year.

**Table S8**: Hazard ratios (HR) comparing mortality (all-cause, natural deaths, unnatural deaths) among individuals with mental health diagnoses to those without unadjusted and adjusted for psychiatric comorbidity.

| **Disorder type** | **Cause of death** | **Sex** | **HR, unadjusted for psychiatric comorbidity** | **HR, adjusted for psychiatric comorbidity** | **p-value for interaction with sex** |
| --- | --- | --- | --- | --- | --- |
| Any Mental Health Diagnosis | All | Both | 1.65 [1.61; 1.69] | - | <0.001 |
|  |  | Female | 1.55 [1.50; 1.61] | - | - |
|  |  | Male | 1.74 [1.68; 1.80] | - | - |
|  | Natural | Both | 1.66 [1.62; 1.70] | - | <0.001 |
|  |  | Female | 1.54 [1.48; 1.60] | - | - |
|  |  | Male | 1.77 [1.71; 1.84] | - | - |
|  | Unnatural | Both | 1.60 [1.48; 1.74] | - | 0.087 |
|  |  | Female | 1.71 [1.46; 2.00] | - | - |
|  |  | Male | 1.56 [1.42; 1.72] | - | - |
| Organic Disorders | All | Both | 6.01 [5.74; 6.29] | 5.04 [4.80; 5.29] | 0.933 |
|  |  | Female | 6.02 [5.63; 6.43] | 5.19 [4.84; 5.57] | - |
|  |  | Male | 6.01 [5.64; 6.40] | 4.90 [4.58; 5.23] | - |
|  | Natural | Both | 6.20 [5.91; 6.50] | 5.30 [5.04; 5.57] | 0.049 |
|  |  | Female | 6.08 [5.67; 6.50] | 5.34 [4.97; 5.74] | - |
|  |  | Male | 6.32 [5.92; 6.75] | 5.25 [4.90; 5.62] | - |
|  | Unnatural | Both | 3.63 [2.79; 4.73] | 2.49 [1.89; 3.27] | 0.020 |
|  |  | Female | 5.13 [3.37; 7.80] | 3.44 [2.23; 5.32] | - |
|  |  | Male | 3.02 [2.14; 4.26] | 2.09 [1.46; 2.97] | - |
| Substance Use Disorders | All | Both | 2.93 [2.64; 3.25] | 1.84 [1.65; 2.05] | 0.247 |
|  |  | Female | 2.63 [2.15; 3.23] | 1.57 [1.28; 1.93] | - |
|  |  | Male | 3.05 [2.70; 3.45] | 1.94 [1.71; 2.20] | - |
|  | Natural | Both | 2.68 [2.37; 3.02] | 1.66 [1.47; 1.87] | 0.307 |
|  |  | Female | 2.42 [1.94; 3.02] | 1.45 [1.16; 1.81] | - |
|  |  | Male | 2.80 [2.43; 3.23] | 1.74 [1.50; 2.01] | - |
|  | Unnatural | Both | 3.64 [2.83; 4.68] | 2.38 [1.83; 3.09] | 0.392 |
|  |  | Female | 4.41 [2.28; 8.51] | 2.49 [1.27; 4.88] | - |
|  |  | Male | 3.53 [2.69; 4.63] | 2.45 [1.84; 3.25] | - |
| Psychotic Disorders | All | Both | 3.12 [2.83; 3.43] | 1.48 [1.34; 1.64] | 0.807 |
|  |  | Female | 3.08 [2.69; 3.54] | 1.56 [1.35; 1.79] | - |
|  |  | Male | 3.15 [2.75; 3.60] | 1.41 [1.23; 1.63] | - |
|  | Natural | Both | 3.13 [2.83; 3.47] | 1.48 [1.33; 1.64] | 0.289 |
|  |  | Female | 3.00 [2.60; 3.48] | 1.52 [1.30; 1.76] | - |
|  |  | Male | 3.27 [2.83; 3.77] | 1.43 [1.23; 1.66] | - |
|  | Unnatural | Both | 2.91 [2.05; 4.12] | 1.57 [1.09; 2.25] | 0.134 |
|  |  | Female | 4.02 [2.27; 7.12] | 2.08 [1.15; 3.76] | - |
|  |  | Male | 2.49 [1.60; 3.88] | 1.38 [0.88; 2.18] | - |
| Mood Disorders | All | Both | 1.59 [1.55; 1.64] | 1.34 [1.30; 1.38] | <0.001 |
|  |  | Female | 1.49 [1.43; 1.56] | 1.28 [1.23; 1.34] | - |
|  |  | Male | 1.70 [1.63; 1.77] | 1.39 [1.34; 1.46] | - |
|  | Natural | Both | 1.56 [1.52; 1.61] | 1.31 [1.26; 1.35] | <0.001 |
|  |  | Female | 1.46 [1.39; 1.52] | 1.25 [1.19; 1.31] | - |
|  |  | Male | 1.69 [1.61; 1.76] | 1.37 [1.31; 1.44] | - |
|  | Unnatural | Both | 1.85 [1.69; 2.04] | 1.58 [1.43; 1.76] | 0.113 |
|  |  | Female | 1.99 [1.68; 2.35] | 1.70 [1.41; 2.03] | - |
|  |  | Male | 1.79 [1.60; 2.01] | 1.53 [1.35; 1.74] | - |
| Anxiety Disorders | All | Both | 1.24 [1.20; 1.27] | 1.03 [0.99; 1.06] | <0.001 |
|  |  | Female | 1.17 [1.12; 1.22] | 0.98 [0.94; 1.03] | - |
|  |  | Male | 1.31 [1.25; 1.36] | 1.07 [1.02; 1.12] | - |
|  | Natural | Both | 1.21 [1.17; 1.25] | 1.01 [0.97; 1.04] | <0.001 |
|  |  | Female | 1.14 [1.09; 1.20] | 0.97 [0.92; 1.01] | - |
|  |  | Male | 1.29 [1.23; 1.35] | 1.05 [1.00; 1.11] | - |
|  | Unnatural | Both | 1.46 [1.32; 1.60] | 1.20 [1.08; 1.33] | 0.111 |
|  |  | Female | 1.56 [1.32; 1.86] | 1.25 [1.04; 1.51] | - |
|  |  | Male | 1.41 [1.26; 1.58] | 1.17 [1.04; 1.33] | - |
| Developmental Disorders | All | Both | 1.67 [1.31; 2.12] | 1.24 [0.97; 1.58] | 0.474 |
|  |  | Female | 1.83 [1.28; 2.62] | 1.41 [0.98; 2.01] | - |
|  |  | Male | 1.55 [1.12; 2.15] | 1.14 [0.82; 1.58] | - |
|  | Natural | Both | 1.94 [1.50; 2.50] | 1.40 [1.08; 1.80] | 0.512 |
|  |  | Female | 2.05 [1.42; 2.95] | 1.56 [1.08; 2.24] | - |
|  |  | Male | 1.84 [1.29; 2.64] | 1.27 [0.88; 1.82] | - |
|  | Unnatural | Both | 0.66 [0.27; 1.59] | 0.54 [0.22; 1.30] | 0.963 |
|  |  | Female | . | . | - |
|  |  | Male | 0.85 [0.35; 2.04] | 0.70 [0.29; 1.70] | - |
| Personality Disorders | All | Both | 1.65 [1.27; 2.15] | 1.00 [0.76; 1.30] | 0.618 |
|  |  | Female | 1.53 [1.01; 2.32] | 0.96 [0.63; 1.46] | - |
|  |  | Male | 1.75 [1.24; 2.46] | 1.03 [0.73; 1.45] | - |
|  | Natural | Both | 1.60 [1.19; 2.13] | 0.96 [0.72; 1.29] | 0.568 |
|  |  | Female | 1.45 [0.93; 2.28] | 0.91 [0.58; 1.42] | - |
|  |  | Male | 1.71 [1.17; 2.50] | 1.01 [0.69; 1.47] | - |
|  | Unnatural | Both | 2.62 [1.36; 5.04] | 1.63 [0.84; 3.14] | 0.594 |
|  |  | Female | 3.26 [1.05; 10.15] | 2.13 [0.68; 6.64] | - |
|  |  | Male | 2.38 [1.07; 5.31] | 1.49 [0.66; 3.33] | - |
| Alcohol Use Disorders | All | Both | 3.22 [2.84; 3.65] | - | 0.427 |
|  |  | Female | 2.93 [2.24; 3.83] | - | - |
|  |  | Male | 3.31 [2.87; 3.81] | - | - |
|  | Natural | Both | 3.01 [2.61; 3.46] | - | 0.616 |
|  |  | Female | 2.84 [2.14; 3.77] | - | - |
|  |  | Male | 3.07 [2.60; 3.61] | - | - |
|  | Unnatural | Both | 4.21 [3.10; 5.71] | - | 0.570 |
|  |  | Female | 4.98 [2.06; 12.02] | - | - |
|  |  | Male | 4.12 [2.97; 5.71] | - | - |
| Drug Use Disorders | All | Both | 2.64 [2.23; 3.11] | - | 0.319 |
|  |  | Female | 2.32 [1.73; 3.11] | - | - |
|  |  | Male | 2.82 [2.30; 3.45] | - | - |
|  | Natural | Both | 2.23 [1.82; 2.73] | - | 0.325 |
|  |  | Female | 1.93 [1.37; 2.70] | - | - |
|  |  | Male | 2.45 [1.90; 3.15] | - | - |
|  | Unnatural | Both | 3.45 [2.40; 4.95] | - | 0.207 |
|  |  | Female | 5.16 [2.31; 11.55] | - | - |
|  |  | Male | 3.18 [2.12; 4.77] | - | - |
| Bipolar Disorders | All | Both | 1.69 [1.57; 1.82] | - | 0.770 |
|  |  | Female | 1.71 [1.55; 1.88] | - | - |
|  |  | Male | 1.67 [1.50; 1.86] | - | - |
|  | Natural | Both | 1.54 [1.42; 1.67] | - | 0.816 |
|  |  | Female | 1.54 [1.38; 1.71] | - | - |
|  |  | Male | 1.55 [1.37; 1.74] | - | - |
|  | Unnatural | Both | 2.77 [2.28; 3.37] | - | 0.103 |
|  |  | Female | 3.26 [2.41; 4.40] | - | - |
|  |  | Male | 2.50 [1.93; 3.23] | - | - |
| Depressive Disorders | All | Both | 1.59 [1.54; 1.63] | - | <0.001 |
|  |  | Female | 1.49 [1.43; 1.55] | - | - |
|  |  | Male | 1.70 [1.63; 1.77] | - | - |
|  | Natural | Both | 1.56 [1.51; 1.61] | - | <0.001 |
|  |  | Female | 1.45 [1.39; 1.52] | - | - |
|  |  | Male | 1.69 [1.61; 1.76] | - | - |
|  | Unnatural | Both | 1.85 [1.68; 2.03] | - | 0.126 |
|  |  | Female | 1.97 [1.67; 2.33] | - | - |
|  |  | Male | 1.79 [1.59; 2.01] | - | - |
| Generalized Anxiety Disorders | All | Both | 1.12 [1.05; 1.19] | - | 0.108 |
|  |  | Female | 1.07 [0.98; 1.16] | - | - |
|  |  | Male | 1.18 [1.08; 1.29] | - | - |
|  | Natural | Both | 1.09 [1.02; 1.16] | - | 0.050 |
|  |  | Female | 1.03 [0.95; 1.13] | - | - |
|  |  | Male | 1.16 [1.05; 1.28] | - | - |
|  | Unnatural | Both | 1.30 [1.06; 1.59] | - | 0.500 |
|  |  | Female | 1.35 [0.97; 1.89] | - | - |
|  |  | Male | 1.27 [0.98; 1.64] | - | - |
| Post-traumatic Stress Disorders | All | Both | 1.23 [1.11; 1.37] | - | 0.449 |
|  |  | Female | 1.19 [1.02; 1.38] | - | - |
|  |  | Male | 1.28 [1.10; 1.49] | - | - |
|  | Natural | Both | 1.20 [1.07; 1.35] | - | 0.750 |
|  |  | Female | 1.17 [1.00; 1.38] | - | - |
|  |  | Male | 1.23 [1.04; 1.45] | - | - |
|  | Unnatural | Both | 1.56 [1.17; 2.07] | - | 0.697 |
|  |  | Female | 1.61 [0.99; 2.62] | - | - |
|  |  | Male | 1.53 [1.08; 2.17] | - | - |
| Eating Disorders | All | Both | 2.52 [1.99; 3.19] | - | 0.899 |
|  |  | Female | 2.56 [1.86; 3.52] | - | - |
|  |  | Male | 2.48 [1.74; 3.53] | - | - |
|  | Natural | Both | 2.65 [2.07; 3.40] | - | 0.386 |
|  |  | Female | 2.39 [1.69; 3.39] | - | - |
|  |  | Male | 2.98 [2.10; 4.24] | - | - |
|  | Unnatural | Both | 1.82 [0.76; 4.38] | - | 0.959 |
|  |  | Female | 4.68 [1.94; 11.29] | - | - |
|  |  | Male | . | - | - |

We adjusted all models for baseline age, and time-updated calendar year. The models combining both sexes were adjusted for sex. There were no deaths from unnatural causes among women with developmental disorders or among men with eating disorders; the corresponding HRs are not shown. Models adjusted for psychiatric comorbidity included binary indicators for organic mental disorders, substance use disorders, psychotic disorders, mood disorders, anxiety disorders, behavioural syndromes associated with physiological disturbances and physical factors, personality disorders, developmental disorders and behavioural disorders.

**Table S9**: Hazard ratios (HR) comparing mortality (all-cause and natural and unnatural deaths) among individuals with mental health diagnoses from hospital settings with those without unadjusted and adjusted for psychiatric comorbidity.

| **Disorder type** | **Cause of death** | **Sex** | **HR, unadjusted for comorbidity** | **HR, adjusted for comorbidity** | **p-value for interaction with sex** |
| --- | --- | --- | --- | --- | --- |
| Any Mental Health Diagnosis | All | Both | 3.79 [3.66; 3.93] | - | <0.001 |
|  |  | Female | 3.42 [3.25; 3.60] | - | - |
|  |  | Male | 4.22 [4.01; 4.44] | - | - |
|  | Natural | Both | 3.87 [3.72; 4.02] | - | <0.001 |
|  |  | Female | 3.41 [3.23; 3.60] | - | - |
|  |  | Male | 4.43 [4.20; 4.68] | - | - |
|  | Unnatural | Both | 3.26 [2.87; 3.72] | - | 0.464 |
|  |  | Female | 3.35 [2.69; 4.17] | - | - |
|  |  | Male | 3.22 [2.75; 3.78] | - | - |
| Organic Disorders | All | Both | 8.48 [8.03; 8.96] | 6.81 [6.43; 7.22] | 0.629 |
|  |  | Female | 8.34 [7.70; 9.03] | 6.91 [6.35; 7.52] | - |
|  |  | Male | 8.61 [7.99; 9.28] | 6.73 [6.22; 7.28] | - |
|  | Natural | Both | 8.77 [8.29; 9.27] | 7.21 [6.80; 7.65] | 0.019 |
|  |  | Female | 8.41 [7.75; 9.13] | 7.13 [6.54; 7.78] | - |
|  |  | Male | 9.10 [8.43; 9.82] | 7.27 [6.70; 7.88] | - |
|  | Unnatural | Both | 4.05 [2.78; 5.91] | 2.50 [1.69; 3.68] | 0.050 |
|  |  | Female | 6.16 [3.43; 11.05] | 3.66 [2.00; 6.72] | - |
|  |  | Male | 3.23 [1.97; 5.31] | 2.02 [1.22; 3.36] | - |
| Substance Use Disorders | All | Both | 3.35 [2.97; 3.77] | 1.97 [1.74; 2.22] | 0.721 |
|  |  | Female | 3.47 [2.70; 4.47] | 1.79 [1.39; 2.31] | - |
|  |  | Male | 3.31 [2.89; 3.80] | 1.99 [1.73; 2.29] | - |
|  | Natural | Both | 3.04 [2.65; 3.50] | 1.75 [1.52; 2.02] | 0.912 |
|  |  | Female | 3.04 [2.29; 4.03] | 1.56 [1.17; 2.08] | - |
|  |  | Male | 3.05 [2.59; 3.58] | 1.78 [1.51; 2.10] | - |
|  | Unnatural | Both | 3.95 [3.01; 5.20] | 2.49 [1.87; 3.30] | 0.095 |
|  |  | Female | 6.51 [3.24; 13.10] | 3.32 [1.62; 6.77] | - |
|  |  | Male | 3.69 [2.74; 4.96] | 2.47 [1.82; 3.37] | - |
| Psychotic Disorders | All | Both | 4.50 [3.90; 5.18] | 1.62 [1.40; 1.88] | 0.804 |
|  |  | Female | 4.58 [3.76; 5.58] | 1.76 [1.44; 2.16] | - |
|  |  | Male | 4.41 [3.60; 5.41] | 1.48 [1.20; 1.83] | - |
|  | Natural | Both | 4.62 [3.97; 5.37] | 1.63 [1.40; 1.91] | 0.677 |
|  |  | Female | 4.51 [3.66; 5.55] | 1.73 [1.40; 2.14] | - |
|  |  | Male | 4.74 [3.81; 5.89] | 1.52 [1.22; 1.90] | - |
|  | Unnatural | Both | 4.39 [2.76; 6.98] | 2.00 [1.24; 3.24] | 0.059 |
|  |  | Female | 7.44 [3.70; 14.97] | 3.17 [1.53; 6.56] | - |
|  |  | Male | 3.30 [1.77; 6.15] | 1.57 [0.83; 2.96] | - |
| Mood Disorders | All | Both | 2.64 [2.52; 2.77] | 1.80 [1.71; 1.90] | <0.001 |
|  |  | Female | 2.44 [2.29; 2.60] | 1.78 [1.66; 1.91] | - |
|  |  | Male | 2.92 [2.72; 3.13] | 1.84 [1.70; 1.99] | - |
|  | Natural | Both | 2.59 [2.47; 2.73] | 1.76 [1.66; 1.86] | <0.001 |
|  |  | Female | 2.37 [2.21; 2.53] | 1.72 [1.59; 1.85] | - |
|  |  | Male | 2.94 [2.73; 3.17] | 1.82 [1.68; 1.98] | - |
|  | Unnatural | Both | 3.07 [2.65; 3.55] | 2.26 [1.91; 2.66] | 0.327 |
|  |  | Female | 3.24 [2.57; 4.09] | 2.45 [1.89; 3.17] | - |
|  |  | Male | 2.96 [2.45; 3.57] | 2.12 [1.71; 2.62] | - |
| Anxiety Disorders | All | Both | 2.80 [2.54; 3.10] | 1.71 [1.54; 1.89] | 0.006 |
|  |  | Female | 2.51 [2.20; 2.86] | 1.61 [1.40; 1.84] | - |
|  |  | Male | 3.33 [2.85; 3.88] | 1.90 [1.62; 2.23] | - |
|  | Natural | Both | 2.73 [2.45; 3.04] | 1.68 [1.50; 1.87] | 0.004 |
|  |  | Female | 2.42 [2.10; 2.79] | 1.57 [1.35; 1.81] | - |
|  |  | Male | 3.32 [2.80; 3.93] | 1.92 [1.61; 2.28] | - |
|  | Unnatural | Both | 3.60 [2.67; 4.86] | 2.09 [1.53; 2.84] | 0.713 |
|  |  | Female | 3.68 [2.35; 5.75] | 2.12 [1.33; 3.37] | - |
|  |  | Male | 3.54 [2.36; 5.30] | 2.00 [1.32; 3.04] | - |
| Developmental Disorders | All | Both | 3.99 [2.15; 7.43] | 2.59 [1.39; 4.82] | 0.398 |
|  |  | Female | 4.91 [2.34; 10.31] | 3.17 [1.51; 6.67] | - |
|  |  | Male | 2.79 [0.90; 8.65] | 1.81 [0.58; 5.61] | - |
|  | Natural | Both | 4.07 [2.12; 7.83] | 2.64 [1.37; 5.09] | 0.721 |
|  |  | Female | 4.47 [2.01; 9.97] | 2.88 [1.29; 6.44] | - |
|  |  | Male | 3.46 [1.11; 10.72] | 2.24 [0.72; 6.95] | - |
|  | Unnatural | Both | . | . | . |
|  |  | Female | . | . | - |
|  |  | Male | . | . | - |
| Personality Disorders | All | Both | 2.79 [1.71; 4.55] | 1.22 [0.75; 2.00] | 0.805 |
|  |  | Female | 2.99 [1.49; 5.97] | 1.87 [0.94; 3.76] | - |
|  |  | Male | 2.61 [1.31; 5.23] | 0.90 [0.45; 1.80] | - |
|  | Natural | Both | 2.25 [1.25; 4.06] | 1.04 [0.57; 1.87] | 0.709 |
|  |  | Female | 2.51 [1.13; 5.58] | 1.61 [0.72; 3.59] | - |
|  |  | Male | 2.00 [0.83; 4.82] | 0.71 [0.30; 1.72] | - |
|  | Unnatural | Both | 7.65 [3.18; 18.39] | 2.80 [1.15; 6.82] | 0.624 |
|  |  | Female | 9.98 [2.49; 40.02] | 4.79 [1.18; 19.46] | - |
|  |  | Male | 6.62 [2.13; 20.54] | 2.31 [0.73; 7.29] | - |
| Alcohol Use Disorders | All | Both | 3.46 [3.00; 3.98] | - | 0.990 |
|  |  | Female | 3.46 [2.48; 4.82] | - | - |
|  |  | Male | 3.46 [2.96; 4.04] | - | - |
|  | Natural | Both | 3.21 [2.74; 3.77] | - | 0.933 |
|  |  | Female | 3.16 [2.20; 4.56] | - | - |
|  |  | Male | 3.23 [2.70; 3.86] | - | - |
|  | Unnatural | Both | 4.36 [3.11; 6.12] | - | 0.126 |
|  |  | Female | 7.89 [3.27; 19.04] | - | - |
|  |  | Male | 4.04 [2.80; 5.84] | - | - |
| Drug Use Disorders | All | Both | 3.34 [2.75; 4.07] | - | 0.859 |
|  |  | Female | 3.43 [2.38; 4.94] | - | - |
|  |  | Male | 3.31 [2.62; 4.19] | - | - |
|  | Natural | Both | 2.88 [2.25; 3.68] | - | 0.987 |
|  |  | Female | 2.78 [1.81; 4.27] | - | - |
|  |  | Male | 2.93 [2.17; 3.95] | - | - |
|  | Unnatural | Both | 3.82 [2.57; 5.68] | - | 0.267 |
|  |  | Female | 6.05 [2.26; 16.18] | - | - |
|  |  | Male | 3.57 [2.32; 5.50] | - | - |
| Bipolar Disorders | All | Both | 2.42 [2.15; 2.71] | - | 0.768 |
|  |  | Female | 2.39 [2.05; 2.77] | - | - |
|  |  | Male | 2.46 [2.06; 2.94] | - | - |
|  | Natural | Both | 2.08 [1.82; 2.37] | - | 0.766 |
|  |  | Female | 2.04 [1.72; 2.43] | - | - |
|  |  | Male | 2.13 [1.73; 2.63] | - | - |
|  | Unnatural | Both | 4.53 [3.47; 5.91] | - | 0.133 |
|  |  | Female | 5.51 [3.74; 8.12] | - | - |
|  |  | Male | 3.90 [2.70; 5.63] | - | - |
| Depressive Disorders | All | Both | 2.65 [2.53; 2.78] | - | <0.001 |
|  |  | Female | 2.46 [2.30; 2.62] | - | - |
|  |  | Male | 2.93 [2.73; 3.15] | - | - |
|  | Natural | Both | 2.62 [2.49; 2.76] | - | <0.001 |
|  |  | Female | 2.40 [2.24; 2.57] | - | - |
|  |  | Male | 2.98 [2.75; 3.22] | - | - |
|  | Unnatural | Both | 2.93 [2.51; 3.42] | - | 0.593 |
|  |  | Female | 2.97 [2.32; 3.81] | - | - |
|  |  | Male | 2.90 [2.38; 3.54] | - | - |
| Generalized Anxiety Disorders | All | Both | 1.51 [1.08; 2.10] | - | 0.396 |
|  |  | Female | 1.68 [1.13; 2.51] | - | - |
|  |  | Male | 1.24 [0.68; 2.23] | - | - |
|  | Natural | Both | 1.52 [1.07; 2.16] | - | 0.675 |
|  |  | Female | 1.61 [1.05; 2.47] | - | - |
|  |  | Male | 1.37 [0.73; 2.54] | - | - |
|  | Unnatural | Both | 1.43 [0.46; 4.45] | - | 0.325 |
|  |  | Female | 2.45 [0.61; 9.82] | - | - |
|  |  | Male | 0.78 [0.11; 5.58] | - | - |
| Post-traumatic Stress Disorders | All | Both | 1.74 [1.25; 2.41] | - | 0.015 |
|  |  | Female | 1.17 [0.71; 1.94] | - | - |
|  |  | Male | 2.65 [1.73; 4.07] | - | - |
|  | Natural | Both | 0.96 [0.60; 1.55] | - | 0.156 |
|  |  | Female | 0.70 [0.35; 1.40] | - | - |
|  |  | Male | 1.43 [0.74; 2.74] | - | - |
|  | Unnatural | Both | 6.68 [4.08; 10.94] | - | 0.895 |
|  |  | Female | 6.73 [3.19; 14.19] | - | - |
|  |  | Male | 6.65 [3.45; 12.81] | - | - |
| Eating Disorders | All | Both | 9.90 [5.97; 16.43] | - | 0.872 |
|  |  | Female | 10.13 [5.75; 17.86] | - | - |
|  |  | Male | 9.07 [2.93; 28.15] | - | - |
|  | Natural | Both | 8.55 [4.73; 15.44] | - | 0.540 |
|  |  | Female | 7.70 [3.85; 15.42] | - | - |
|  |  | Male | 12.05 [3.88; 37.38] | - | - |
|  | Unnatural | Both | 17.20 [5.54; 53.45] | - | 0.973 |
|  |  | Female | 28.69 [9.21; 89.39] | - | - |
|  |  | Male | . | - | - |

We adjusted all models for baseline age, and time-updated calendar year. We adjusted the models combining both sexes for sex. There were no deaths from unnatural causes among men with developmental disorders or eating disorders, or among people of either sex with developmental disorders; corresponding HR are not shown. Models adjusted for psychiatric comorbidity included binary indicators for organic mental disorders, substance use disorders, psychotic disorders, mood disorders, anxiety disorders, behavioural syndromes associated with physiological disturbances and physical factors, personality disorders, developmental disorders and behavioural disorders.

**Table S10**: Hazard ratios (HR) comparing mortality (all-cause, natural deaths, unnatural deaths) among individuals prescribed psychiatric medication to those who were not prescribed psychiatric medication unadjusted and adjusted for psychiatric comorbidity.

| **Medication type** | **Cause of death** | **Sex** | **HR, unadjusted for comorbidity** | **HR, adjusted for comorbidity** | **p-value for interaction with sex** |
| --- | --- | --- | --- | --- | --- |
| Any Medication | All | Both | 1.77 [1.73; 1.81] | - | <0.001 |
|  |  | Female | 1.64 [1.58; 1.70] | - | - |
|  |  | Male | 1.87 [1.81; 1.93] | - | - |
|  | Natural | Both | 1.84 [1.79; 1.88] | - | <0.001 |
|  |  | Female | 1.66 [1.59; 1.72] | - | - |
|  |  | Male | 1.99 [1.92; 2.06] | - | - |
|  | Unnatural | Both | 1.37 [1.26; 1.48] | - | 0.537 |
|  |  | Female | 1.33 [1.14; 1.56] | - | - |
|  |  | Male | 1.38 [1.25; 1.51] | - | - |
| Antidepressants | All | Both | 1.63 [1.59; 1.67] | 1.41 [1.37; 1.45] | <0.001 |
|  |  | Female | 1.51 [1.46; 1.57] | 1.33 [1.28; 1.38] | - |
|  |  | Male | 1.74 [1.68; 1.79] | 1.48 [1.43; 1.53] | - |
|  | Natural | Both | 1.64 [1.60; 1.69] | 1.41 [1.37; 1.45] | <0.001 |
|  |  | Female | 1.50 [1.45; 1.56] | 1.32 [1.27; 1.38] | - |
|  |  | Male | 1.78 [1.72; 1.85] | 1.50 [1.44; 1.56] | - |
|  | Unnatural | Both | 1.52 [1.40; 1.66] | 1.41 [1.29; 1.55] | 0.662 |
|  |  | Female | 1.49 [1.27; 1.75] | 1.32 [1.11; 1.56] | - |
|  |  | Male | 1.54 [1.39; 1.70] | 1.45 [1.30; 1.62] | - |
| Antipsychotics | All | Both | 2.18 [2.12; 2.24] | 1.93 [1.87; 1.99] | <0.001 |
|  |  | Female | 2.00 [1.92; 2.09] | 1.81 [1.73; 1.89] | - |
|  |  | Male | 2.36 [2.27; 2.46] | 2.05 [1.97; 2.14] | - |
|  | Natural | Both | 2.24 [2.17; 2.31] | 1.98 [1.92; 2.04] | <0.001 |
|  |  | Female | 2.01 [1.93; 2.10] | 1.83 [1.75; 1.91] | - |
|  |  | Male | 2.49 [2.39; 2.60] | 2.15 [2.05; 2.24] | - |
|  | Unnatural | Both | 1.56 [1.39; 1.75] | 1.39 [1.24; 1.57] | 0.082 |
|  |  | Female | 1.70 [1.41; 2.07] | 1.53 [1.25; 1.86] | - |
|  |  | Male | 1.48 [1.28; 1.71] | 1.32 [1.14; 1.53] | - |
| Anxiolytics | All | Both | 1.44 [1.40; 1.47] | 1.18 [1.15; 1.21] | <0.001 |
|  |  | Female | 1.36 [1.31; 1.41] | 1.13 [1.09; 1.18] | - |
|  |  | Male | 1.51 [1.46; 1.56] | 1.23 [1.18; 1.27] | - |
|  | Natural | Both | 1.45 [1.42; 1.49] | 1.19 [1.16; 1.22] | <0.001 |
|  |  | Female | 1.35 [1.30; 1.41] | 1.13 [1.08; 1.18] | - |
|  |  | Male | 1.56 [1.50; 1.61] | 1.25 [1.20; 1.30] | - |
|  | Unnatural | Both | 1.30 [1.19; 1.42] | 1.13 [1.03; 1.24] | 0.080 |
|  |  | Female | 1.40 [1.19; 1.65] | 1.21 [1.01; 1.44] | - |
|  |  | Male | 1.26 [1.13; 1.40] | 1.10 [0.98; 1.23] | - |
| Substance Use Medication | All | Both | 1.29 [1.11; 1.50] | 1.11 [0.95; 1.29] | 0.033 |
|  |  | Female | 1.58 [1.25; 2.00] | 1.33 [1.05; 1.68] | - |
|  |  | Male | 1.14 [0.94; 1.39] | 0.99 [0.81; 1.21] | - |
|  | Natural | Both | 1.32 [1.12; 1.55] | 1.13 [0.96; 1.32] | 0.043 |
|  |  | Female | 1.61 [1.27; 2.06] | 1.36 [1.06; 1.73] | - |
|  |  | Male | 1.15 [0.93; 1.42] | 0.99 [0.80; 1.23] | - |
|  | Unnatural | Both | 1.04 [0.62; 1.73] | 0.94 [0.57; 1.56] | 0.751 |
|  |  | Female | 1.15 [0.37; 3.58] | 0.98 [0.32; 3.06] | - |
|  |  | Male | 1.01 [0.57; 1.79] | 0.93 [0.53; 1.64] | - |

All models were adjusted for baseline age and time-updated calendar year. The models combining both sexes were adjusted for sex.
